# Increasing risks of recurrence following community-associated *Clostridioides difficile* infection in Oxfordshire 2000-2025

**DOI:** 10.64898/2026.09.30.26364395

**Authors:** Elisabeth Dietz, Julie Robotham, Mark Wilcox, Colin Brown, Russell Hope, Berit Muller-Pebody, David W. Eyre, A. Sarah Walker

## Abstract

**Background:** *Clostridioides difficile* infections (CDI) continue to cause substantial morbidity and mortality, with increasing incidence in some countries post-COVID-19. Recurrence affects 10-20% of CDI patients: whether risk factors have changed over time is unclear, particularly post-COVID-19.

**Methods:** Using electronic health records from hospitals in Oxfordshire, UK, including results from community samples, we estimated the incidence of CDI recurrence across different calendar periods between 2000-2025. To investigate whether risk factors had changed over time, we used interaction tests within multivariable Fine-Gray regression models (modelling death as a competing risk), using backwards elimination, starting from 39 risk factors identified from the literature.

**Results:** Recurrence occurred within 1-year in 12.6%, 12.5%, 10.8% and 13.1% of CDI cases in “high virulence” 2003-2008 (n=527), “post-high virulence” 2008-2012 (n=252), “pre-pandemic” 2012-2020 (n=143) and “post-pandemic” 2020-2025 (n=110) periods respectively. The only factor with evidence of variation in its association with recurrence over time was surveillance classification (heterogeneity p=0.019; others p>0.05). Compared with hospital-onset cases, community-onset community-associated cases had increasing recurrence risks over time (subdistribution hazard ratios (SHRs)=2.05 (95% CI 1.27-3.31) in “post-pandemic” and 1.86 (1.16-2.98) in “pre-pandemic”), especially compared to the “high virulence” era of the early 2000s (SHR=0.81 (0.56-1.16)). This heterogeneity persisted after adjusting for baseline blood tests as a marker of severity (heterogeneity p=0.022).

**Conclusions:** Compared with hospital-onset hospital-associated cases, recurrence risks increased for community-onset community-associated CDI cases over time. Recurrence is a key contributor to overall CDI incidence and the underlying causes of increasing risks with community-onset community-associated cases should be explored further.

**Short summary:** Over 2000-2025, risk of recurrence following *Clostridioides difficile* infection increased significantly for community-onset community-associated cases, compared to hospital-onset hospital-associated cases. Older age, hospital exposure, immunosuppression, chronic kidney disease, diverticular disease, enteral feeding and recent sepsis/bacteraemia were significantly associated with recurrence.

## Background

*Clostridioides difficile* infections (CDI) continue to cause substantial morbidity and mortality globally^1,2^, affecting ∼500,000 patients annually in the USA^3^. In the early 2000s, many countries saw marked rises in CDI incidence, particularly in hospitals, associated with the highly virulent ribotype027 strain^4,5^. This trend was reversed following strengthened hospital infection control measures and reduced prescribing of high-risk antibiotics^6-8^. However, CDI incidence has increased in several countries including England^9^, Spain^10^, and the USA^11^ since COVID-19, for reasons not fully understood. This increase is concerning given the burden CDI places on patients^12,13^ and healthcare systems^14^.

Increased recurrence is a key consequence of rising CDI incidence, as well as a contributor, typically affecting 10-20% of patients^1,15^. Previous studies have identified key risk factors for CDI recurrence as older age and antibiotics^16^, along with prior CDI and hospital exposure^17^, severe initial CDI^18^, and comorbidity burden^19^. Elevated risks have also been reported among specific patient groups, including immunocompromised individuals^20^, transplant recipients^21^, and patients with kidney disease^22^, liver disease^3^, cancer^23^, diverticular disease^24^, and inflammatory bowel disease (IBD)^25^. However, findings are not consistent^3,17^: some studies have been limited by small sample sizes and incomplete confounding adjustment^16,26^, and others have not formally accounted for death as a competing risk, highly pertinent since CDI commonly affects elderly and frail populations^21,27,28^ and all-cause mortality within a year of CDI is high^18^. Risk factors for CDI recurrence post COVID-19 have been rarely studied.

To investigate whether risk factors for CDI recurrence have changed post-COVID, potentially affecting incidence, we analysed Oxfordshire electronic health records spanning more than two decades. Accounting for the competing risk of death, we estimated recurrence risks across different calendar periods, and investigated whether the effect of risk factors for CDI recurrence varied over time.

## Methods

This study analysed de-identified electronic health records from the Infections in Oxfordshire Research Database (IORD) which contains data from Oxford University Hospitals (OUH). OUH comprises four teaching hospitals, and is the sole provider of specialist services and microbiology in the region, serving ∼900,000 people (estimated in 2024^29^). The database includes hospital records and laboratory/microbiology results from samples sent from the community (including primary care, community hospitals, and nursing homes) for testing at OUH laboratories. IORD has Research Ethics Committee, Health Research Authority, and Confidentiality Advisory Group approvals (24/SC/0241, 19/CAG/0144).

### Population

Adults (≥18 years) were followed from each toxin antigen-positive test between April 1^st^ 2000 and March 16^th^ 2025 (de-duplicated using a 28-day window, given laboratory guidelines discouraging repeat testing in recent positives^30^) until recurrence (next toxin-positive test), death, or censoring (end of calendar period or last OUH contact). Mortality was ascertained using hospital and national mortality registry records (excluding 61 patients with inconsistent mortality and healthcare contact records, due to risk of mislinkage of hospital records). CDI diagnosis used a toxin enzyme-linked immunosorbent assay (ELISA) before April 2012 and a two-step algorithm after April 2012 (glutamate dehydrogenase (GDH) ELISA followed by toxin ELISA for GDH-positive samples), defining GDH-positive, toxin-negative samples as carriage. We defined four calendar periods (**Tables 1, S1**) based on testing and positivity trends (**Figures S1-S3**) and national CDI epidemiology.

**Table 1:** Study periods and outcomes.

| Period name | Date range | CDI cases | Patients | One-year cumulative incidence of recurrence | One-year cumulative incidence of death before recurrence |
| --- | --- | --- | --- | --- | --- |
| *Overall study* | 01 April 2000 – 16 March 2025 | 9,456 | 8,046 | 13.0% | 40.7% |
| *High virulence* | 01 April 2003 – 31 March 2008 | 4,324 | 3,737 | 12.6% | 43.2% |
| *Post-high virulence* | 01 April 2008 – 31 March 2012 | 2,065 | 1,798 | 12.5% | 32.5% |
| *Pre-pandemic* | 01 April 2012 – 29 February 2020 | 1,351 | 1,196 | 10.8% | 30.9% |
| *Post-Pandemic* | 01 June 2020 – 16 March 2025 | 881 | 764 | 13.1% | 32.4% |

NOTE: Cumulative incidence was estimated by Aalen-Johansen 365 days after index CDI, using recurrence/death before recurrence as a competing risk. Three months between March and May 2020 were excluded from the post-pandemic period due to substantial healthcare delivery disruption when the SARS-CoV-2 pandemic started. Delayed entry was permitted for those with a positive test in the 365 days prior to period start (last 365 days of the “pre-pandemic” period for delayed entry to “post-pandemic”), further details in Supplementary Table 1. Multiple recurrences per patient permitted, and a small number of patients were excluded due to missing information on sex and/or address.

### Factors

We studied 39 factors identified from the literature (**Tables 2, S2**), including CDI onset/association (surveillance classification). Hospital-onset hospital-associated (HOHA) samples were collected >48h from admission^31^. Community-onset hospital-associated (COHA), community-onset indeterminate-association (COIA) and community-onset community-associated (COCA) samples were collected in the community or within ≤48h of admission, from patients last discharged within ≤4, 4-≤12 or >12 weeks, respectively. We also considered (definitions in **Supplementary Methods**): demographics (age, sex, deprivation and catchment), prior test result history, comorbidity scores (Charlson, Elixhauser, and Gilbert’s Hospital Frailty Risk Score), hospital exposures (emergency and elective bed-days, number of emergency admissions, number of outpatient appointments, and recent surgical and gastrointestinal admission), chronic conditions (palliative care, immunosuppression, cancer, dementia, diabetes, anaemia, kidney disease, liver disease, heart failure, hypertension, other heart disease, cerebrovascular disease, chronic-obstructive pulmonary disease [COPD], IBD, diverticular disease and oesophageal disease), acute/recent conditions (acute renal failure, pneumonia/bronchitis, sepsis/bacteraemia and urinary tract infection [UTI]), and procedure history (transplant receipt and enteral tube feeding).

**Table 2:** Key characteristics of CDI episodes by study period.

| Factor |  | Overall<br>2000-2025 | “High virulence”<br>2003-2008 | “Post-high virulence”<br>2008-2012 | “Pre-pandemic”<br>2012-2020 | “Post-pandemic”<br>2020-2025 |
| --- | --- | --- | --- | --- | --- | --- |
| Surveillance classification (n, %) | HOHA | 5,644 (59.7%) | 2,893 (66.9%) | 1,035 (50.1%) | 550 (40.7%) | 380 (43.1%) |
|  | COHA | 2,001 (21.2%) | 860 (19.9%) | 420 (20.3%) | 395 (29.2%) | 251 (28.5%) |
|  | COIA | 661 (7.0%) | 226 (5.2%) | 199 (9.6%) | 154 (11.4%) | 88 (10.0%) |
|  | COCA | 1,150 (12.2%) | 345 (8.0%) | 411 (19.9%) | 252 (18.7%) | 162 (18.4%) |
| Age (median, IQR) |  | 79.0 (68.0, 85.8) | 79.0 (68.0, 85.8) | 79.4 (69.9, 85.8) | 77.8 (66.1, 85.5) | 78.6 (65.1, 86.5) |
| Sex (n, %) | Male | 4,086 (43.2%) | 1,942 (44.9%) | 872 (42.2%) | 540 (40.0%) | 366 (41.5%) |
|  | Female | 5,370 (56.8%) | 2,382 (55.1%) | 1,193 (57.8%) | 811 (60.0%) | 515 (58.5%) |
| Catchment percentage (median, IQR) |  | 95.4 (90.3, 96.4) | 95.6 (90.6–96.5) | 95.3 (88.3–96.2) | 95.3 (90.6–96.4) | 94.0 (87.5–96.1) |
| Catchment percentage ≥80% (n, %) |  | 7,607 (80.4%) | 3,487 (80.6%) | 1,625 (78.7%) | 1,107 (81.9%) | 707 (80.2%) |
| Deprivation percentile (median, IQR) |  | 76 (57, 89) | 77 (57, 89) | 79 (57, 90) | 73 (52, 88) | 75 (55, 88) |
| Prior test result in past 6 months<br>(n, %) | CDI | 1114 (11.8%) | 521 (12.0%) | 287 (13.9%) | 141 (10.4%) | 104 (11.8%) |
|  | Negative | 3043 (32.2%) | 1493 (34.5%) | 941 (45.6%) | 258 (19.1%) | 203 (23.0%) |
|  | Carriage | — | — | — | 134 (9.9%) | 98 (11.1%) |
| Charlson comorbidity index (median, IQR) |  | 1 (0, 2) | 1 (0, 2) | 1 (0, 2) | 2 (0, 3) | 2 (0, 4) |
| Gilbert frailty score (median, IQR) |  | 3.2 (0.0, 8.4) | 2.4 (0.0, 6.1) | 2.9 (0.0, 8.0) | 7.3 (1.1, 16.0) | 13.1 (4.6, 21.7) |
| Emergency bed-days (weeks in past 12 months) (median, IQR) |  | 0.6 (0, 2.9) | 0.6 (0, 2.8) | 0.3 (0, 2.8) | 0.7 (0, 3.4) | 0.9 (0–3.3) |
| Number of emergency admissions (past 6 months) (median, IQR) |  | 1 (0, 1) | 1 (0, 1) | 0 (0, 1) | 1 (0, 2) | 1 (0, 2) |
| Number of outpatient appointments (past 6 months) (median, IQR) |  | 1 (0, 3) | 1 (0, 3) | 1 (0, 3) | 2 (0, 5) | 2 (0, 6) |
| Immunosuppression (past 5 years) (n, %) |  | 883 (9.3%) | 307 (7.1%) | 186 (9.0%) | 172 (12.7%) | 164 (18.6%) |
| Chronic kidney disease (past 5 years) (n, %) |  | 1,188 (12.6%) | 341 (7.9%) | 227 (11.0%) | 330 (24.4%) | 257 (29.2%) |
| Diverticular disease (past 5 years) (n, %) |  | 1,080 (11.4%) | 384 (8.9%) | 267 (12.9%) | 256 (18.9%) | 182 (20.7%) |
| Sepsis/bacteraemia (past 3 months) (n, %) |  | 641 (6.8%) | 158 (3.7%) | 91 (4.4%) | 167 (12.4%) | 202 (22.9%) |
| Feeding tubes (past 5 years) (n, %) |  | 324 (3.4%) | 131 (3.0%) | 83 (4.0%) | 44 (3.3%) | 68 (7.7%) |
| Neutrophils (x10<sup>9</sup>/L) (median, IQR) |  | 9.8 (6.1, 15.0) | 10.1 (6.4, 15.4) | 9.1 (5.7, 13.8) | 10.1 (6.3, 16.1) | 8.7 (5.4, 13.8) |
| CRP (mg/L) (median, IQR) |  | 96 (43, 160) | 97 (44, 160) | 96 (39, 160) | 96 (43, 156) | 93 (38, 159) |

NOTE: There was ~30% missing data for blood test results, see Supplementary Table S2. Other risk factors investigated in the study are also given in Supplementary Table S2. Deprivation percentile is defined by the Index of Multiple Deprivation: higher values indicate less deprivation. Variables truncated in modelling are here described pre-truncation.

### Statistical analysis

For each toxin-positive test (i.e. including recurrences after 28 days as new episodes), the cumulative incidence of subsequent recurrence, and death before recurrence, was calculated using the nonparametric Aalen-Johansen estimator (R *cmprsk* package^32^), restricted to patients living in areas with ≥80% OUH catchment to minimise ascertainment bias. Subdistribution hazards for recurrence and death before recurrence were modelled using Fine-Gray competing risk regression, restricted to complete cases (excluding 1% with missing address in whom catchment and deprivation could not be calculated), and fitted separately for each outcome (recurrence, death before recurrence) and study period (8 models) using robust standard errors clustered by patient (R *coxph()* function on weighted data generated by the *finegray()* function from the *survival* package^33,34^). Backwards selection with a liberal exit threshold of p>0.1 was performed on each model, to augment confounding adjustment. To test for variation in the effect of risk factors over time, variables selected by at least two models were included in a single stacked Fine-Gray model for recurrence, stratified by period to allow period-specific baseline hazards, and including interaction terms between all covariates and study period. These interaction terms were then pruned using backward selection (exit interaction-p>0.05), to identify evidence of temporal heterogeneity, retaining all main effects regardless of significance, before backward selection of main effects (exit p>0.05) to retain factors with evidence of association with recurrence over the full study period. To test for effects of index case severity, baseline blood test results were then added to this selected model. Sensitivity analyses additionally adjusted for admission-related variables, and used a generalised additive model (GAM) of recurrence over the full study period to estimate when changes in recurrence risk occurred more precisely (**Supplementary Methods**).

## Results

### Recurrence rates and patient characteristics

There were 4,324, 2,065, 1,351 and 881 positive CDI tests in the “high virulence” (2003-2008), “late ELISA” (2008-2012), “pre-pandemic” (2012-2020) and “post-pandemic” (2020-2025) periods respectively, with one-year cumulative incidences of recurrence varying minimally at 12.6%, 12.5%, 10.8% and 13.1% respectively (**Table 1**). The cumulative incidence of death before recurrence varied more, from 43.2% in the “high virulence” period to between 30.9% and 32.5% thereafter. In those with recurrences, time to recurrence was relatively stable across periods (median ∼55 days) (**Table S1**). As expected, positive tests (both first episodes and recurrences) occurred in older individuals (overall median 79 years) (**Table 2**). Hospital exposures and comorbidities were common and generally increased over time, e.g., frailty scores (median 2.4 in “high virulence” vs 13.2 in “postpandemic”) and conditions such as immunosuppression, chronic kidney disease and recent sepsis/bacteraemia (**Table 2)**. The fraction of cases classified as HOHA declined over time: dropping from 67% in “high virulence” to 43% in “post-pandemic” (**Table 2**), with a correspondingly lower fraction of COCA cases in the “high virulence” period.

### Recurrence and mortality by initial surveillance classification

Recurrence after COCA cases in the “high virulence” period was notably lower, compared to the other surveillance classifications (**Figure 1**). In contrast, during the “pre-pandemic” and “postpandemic” periods, recurrence incidence after COCA was similar to COIA and COHA, and higher than after HOHA. The particularly high observed recurrence incidence amongst COIA cases “post-high virulence” is likely due to small numbers.

**Figure 1:**
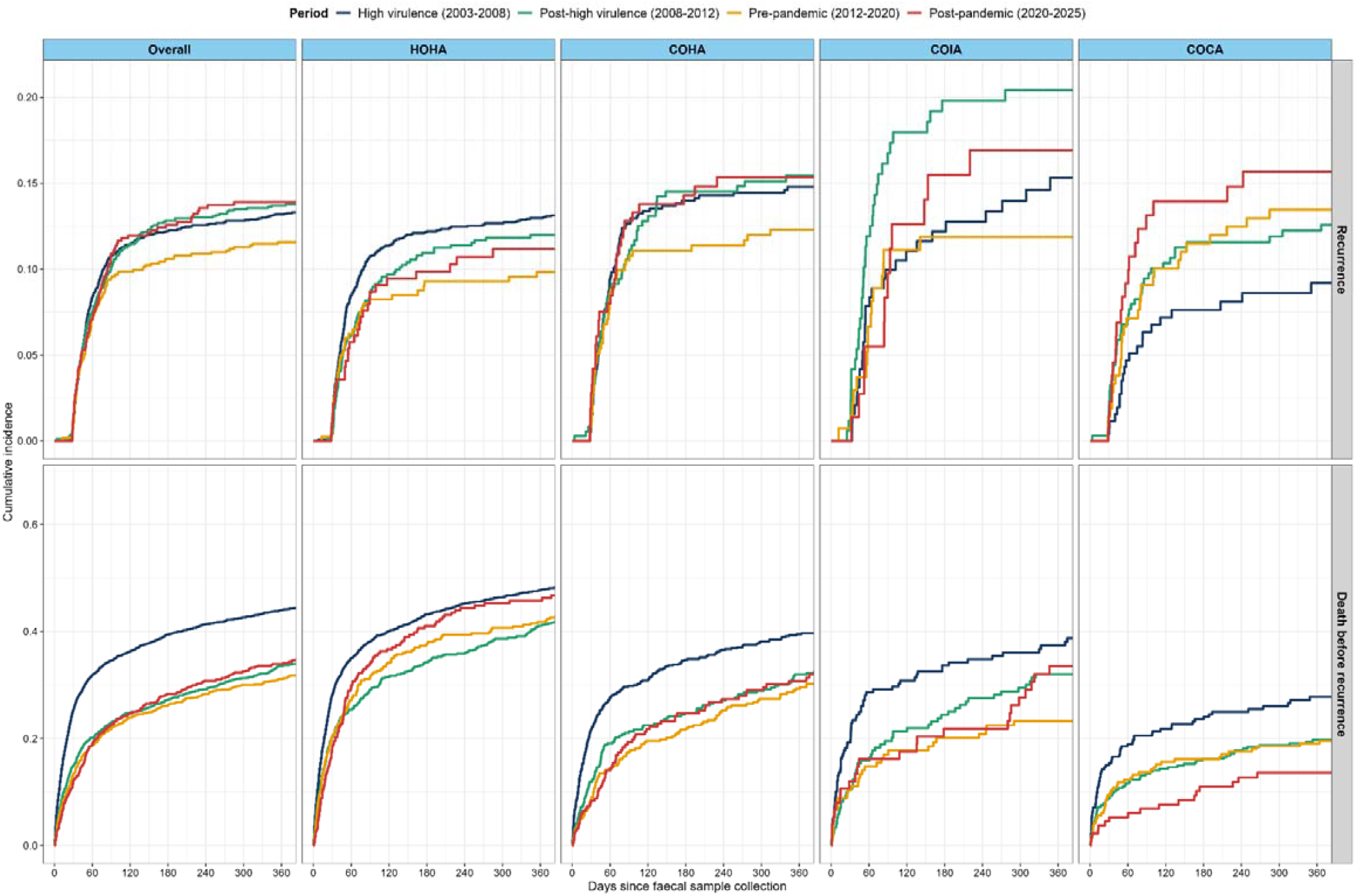
Cumulative incidence of recurrence and death before recurrence, overall and by surveillance classification across time. NOTE: Follow-up truncated at 365 days; Figure S5 shows follow-up three years post-index episode. Estimates by Aalen-Johansen with the other outcome as a competing risk, analysis restricted to patients with OUH catchment ≥80% to minimise ascertainment bias. Gray’s test for differences in cumulative incidence by period were 0.166 (recurrence) and <0.001 (death) overall, 0.049 (recurrence) and <0.001 (death) amongst HOHA cases, 0.648 (recurrence) and <0.001(death) amongst COHA cases, 0.343 (recurrence) and 0.007 (death) amongst COIA cases, and 0.503 (recurrence) and 0.042 (death) amongst COCA cases. Gray’s test for differences in cumulative incidence by surveillance classification were 0.041 (recurrence) and <0.001 (death) pooled, 0.199 (recurrence) and <0.001 (death) in the “high virulence” period, 0.028 (recurrence) and <0.001 (death) in “post-high virulence”, 0.383 (recurrence) and <0.001 (death) in “pre-pandemic”, and 0.393 (recurrence) and <0.001 (death) in “post-pandemic”.

Mortality before recurrence was generally highest for cases with more recent healthcare contact; being highest in HOHA and lowest in COCA consistently across periods. While cumulative mortality before recurrence was consistently higher in the “high virulence” period, the difference between periods for HOHA cases was smaller than expected. One-year COCA mortality was particularly low in the “postpandemic” period, but extending the follow-up period to the first three years post-CDI revealed similar trends to the “pre-pandemic” and “post-high virulence” periods (**Figure S4**).

Comparing surveillance classification of index and recurrent CDI (**Figure 2**) showed increases in community-associated and indeterminant first cases and recurrences after the first “high virulence” period, when nearly half of recurrences were HOHA at both index and recurrence, and another third were HOHA index/COHA recurrence. Similar patterns were seen in each subsequent time period.

**Figure 2:**
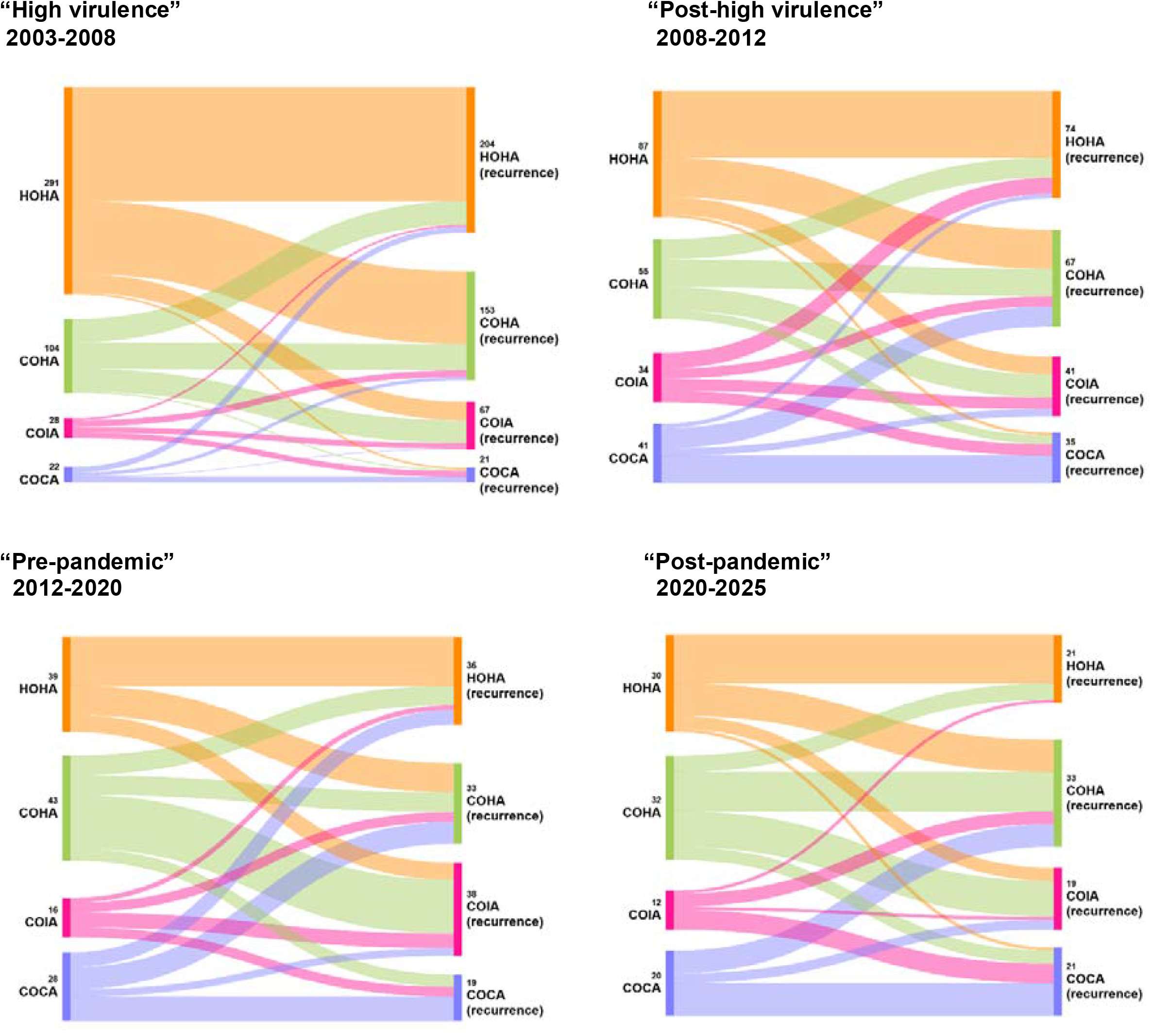
Surveillance classification of C. difficile index cases and recurrences by study period. NOTE: Multiple recurrences per patient permitted. Analysis restricted to patients with OUH catchment ≥80% to minimise ascertainment bias, and to recurrences within 365 days of the index episode.

### Risk factors for recurrence and death before recurrence

We identified an extensive list of candidate risk factors consistently found in period-specific models of recurrence and death before recurrence (**Table S3**), which were combined in a single recurrence model stratified by study period (**Figure S6**). After testing which risk factors had evidence of different effects in different periods (interaction p<0.05), we found that only the effect of surveillance classification on recurrence varied over time (**Figure S7**). We identified multiple risk factors for recurrence across the whole study period, including age, catchment percentage, Charlson index, emergency bed-days, number of emergency admissions, number of outpatient appointments, immunosuppression, chronic kidney disease, diverticular disease, sepsis/bacteraemia, and enteral feeding tubes (**Figure 3**). Excepting the Charlson index, these factors were all associated with increased recurrence risk. The negative association observed for Charlson was likely driven by strong competing risks from death, as evidenced by positive cause-specific associations with death before recurrence and no evidence of cause-specific association with recurrence (**Figure S8**). Significantly lower risk of death before recurrence was associated with other surveillance classifications vs HOHA, whereas older age, and higher Charlson index, number of emergency admissions and outpatient appointments, and immunosuppression were associated with higher mortality risk.

**Figure 3.**
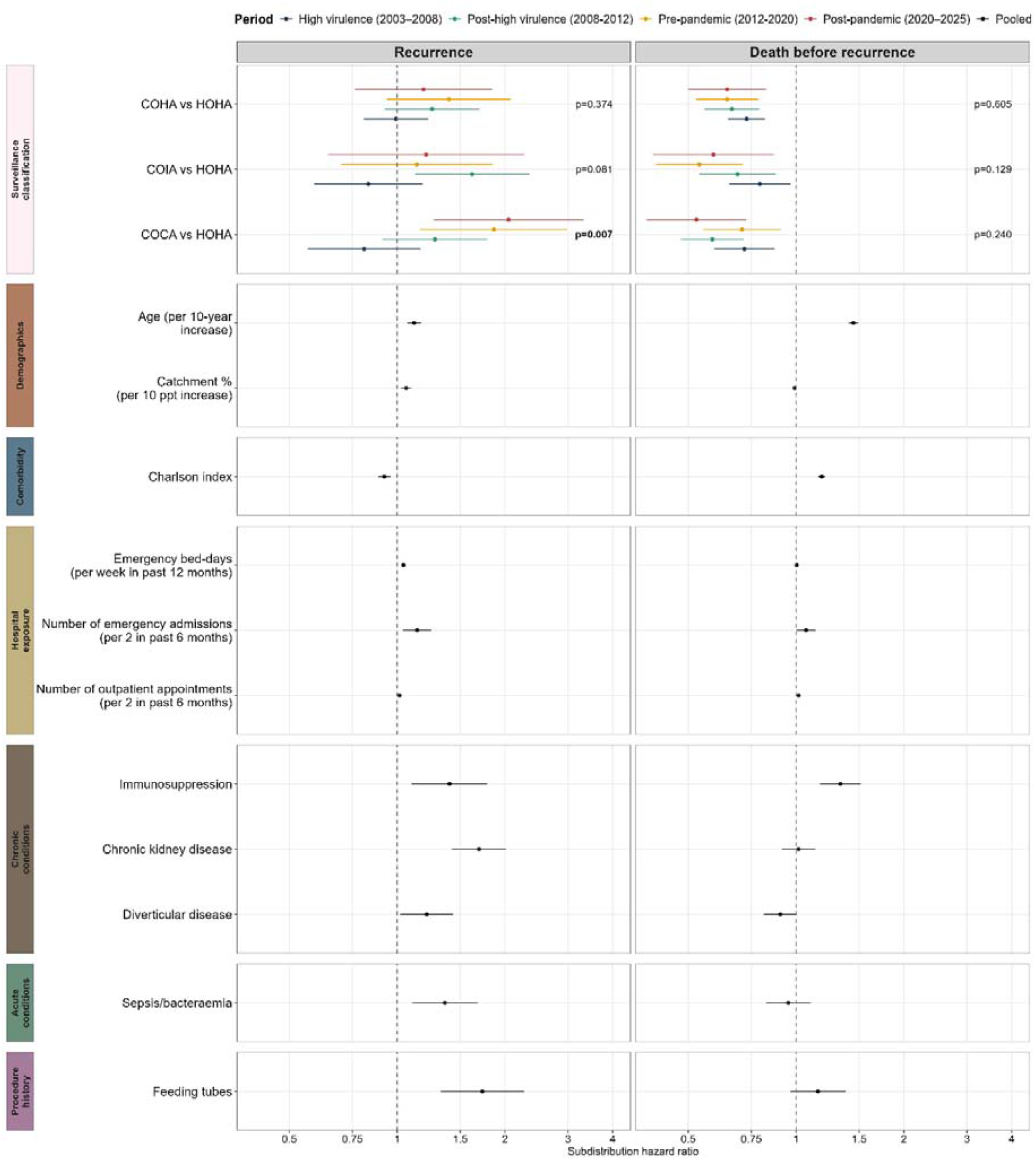
Risk factors for recurrence and death before recurrence. Note: Estimates from Fine-Gray competing risk regression, treating the other outcome as a competing risk. Overall heterogeneity p-values for surveillance classification were 0.019 (recurrence) and 0.273 (death). Models were stratified by period, and robust standard errors were clustered by patient. Pooled variables selected by backward selection from recurrence model displayed in Figure S7. SHRs (95% CI) for recurrence for COCA vs HOHA were 0.81 (0.56, 1.16) in “high virulence”, 1.28 (0.91, 1.79) “post-high virulence”, 1.86 (1.16, 2.98) “prepandemic”, and 2.05 (1.27, 3.31) “post-pandemic”.

Adjusting for other factors, the recurrence risk after a COCA infection was significantly elevated vs HOHA in the last two periods spanning 2012-2025 (heterogeneity p=0.007; **Figure 3**). Period-level effects therefore suggested that risk started increasing before COVID-19. To investigate more closely when this risk changed, using an alternative modelling approach allowing the effect of calendar time to vary smoothly across the whole study period (**Figure S9**), revealed a gradual increase in risk relative to HOHA for all three community-onset categories from ∼2008 to ∼2014 (**Figure S10**), before stabilising and perhaps slightly declining at the end of the study (although with substantial uncertainty). For mortality, there was no evidence of difference across periods in the effect of surveillance definitions (heterogeneity p=0.240) (**Figure 3**).

Cause-specific estimates produced similar findings, with significant heterogeneity between periods in the effect of COCA vs HOHA on recurrence (p=0.008), but not death before recurrence (p=0.234) (**Figure S8**). When restricting follow-up to 365 and 90 days after CDI, significantly elevated recurrence risks for COCA vs HOHA persisted in the “post-pandemic” period (recurrence heterogeneity p=0.005, p=0.011 respectively), but there was also evidence of heterogeneity in mortality (p=0.032, p=0.018) (**Figures S12-S13**), potentially reflecting competing risks given the lower mortality risk in “post-pandemic” for COCA vs HOHA, particularly compared to “high virulence”. Using the same 365- and 90-day follow-up restrictions in cause-specific models, there was consistent evidence for heterogeneity with recurrence (p=0.007 and p=0.013 respectively), and also strengthening evidence with shorter follow-up periods for heterogeneity with mortality (p=0.068, p=0.029) (**Figure S14, Figure S15**). Together, these results suggest that lower mortality among COCA cases may partly have contributed to elevated recurrence risks in the “post-pandemic” period, particularly within 90 days of CDI, but it is unlikely to fully explain the association over longer follow-up.

There was no evidence of association with recurrence for any baseline blood test result, and including these variables did not change the observed effects of surveillance classification (**Figure S16**), suggesting variation over time cannot be easily explained by severity. We observed expected associations between mortality risk and lower albumin, eosinophils, and platelets, and higher neutrophils, creatinine, and CRP. Cause-specific effects suggest that competing risks may affect some associations with blood test results: significant cause-specific associations with recurrence were observed for lower albumin and higher neutrophils and CRP (**Figure S17**), but not when accounting for the competing risk of death before recurrence.

Adjusting for additional admission variables also did not alter surveillance classification heterogeneity (**Figure S18**). As expected, admission within the next 48h was associated with higher risk of both recurrence and death (likely linked to severity), and those without prior OUH admissions had lower mortality risk.

Investigating the COCA cases in more detail, there was no evidence that they were dominated by samples taken in community hospitals, which made up around 5% of total cases in the last two periods (**Figure S19, Table S4**), a decrease from 7.8% in the “high virulence” period, reflecting broader shifts away from this care modality in England. There was notable year-on-year variation in sample location for COCA cases, but the fraction taken in primary care increased steadily from the early 2000s, generally reaching 50% or more in 2008 onwards (**Figure S19**). Cases where samples were taken in primary care were rare in “high virulence” (9%) versus later periods (20.5%-25.2%), and the proportion of non-HOHA hospital samples (within first 48h of an admission) increased from 16.2% in “high virulence” to 31.0% “post-pandemic”.

## Discussion

We have shown that associations between surveillance classification of index cases and CDI recurrence risk have changed significantly over the last 25 years, with COCA cases now at significantly higher risk of recurrence compared with HOHA cases. Whilst we found no evidence that other risk factors for CDI recurrence changed over time, we confirmed several risk factors for recurrence over the full study period. As in previous studies, risk increased with age and prior hospitalisation^1,3,16,17^. Beyond emergency bed-days in the past year, the number of emergency admissions in the past six months also contributed additional risk. This may be due to recency effects, or multiple admissions arising from re-admissions or multimorbidity, indicative of patient frailty. A small but significant positive association was also seen for outpatient appointments. We observed increased recurrence risks for patients with a history of several conditions, notably the well-established factors of immunosuppression and chronic kidney disease^20,22,35,36^. Interestingly, we also observed elevated risks for enteral feeding tube history, which could be caused by altered gut microbiota, but also through associations with prolonged hospitalisation and antibiotic use^37^. It is also possible that diarrhoea associated with enteral feeding may lead to false ascertainment of CDI in colonised patients. The same explanation may also apply to the observed risks for diverticular disease, but this may also be associated with repeated antibiotic exposures. Recent sepsis/bacteraemia was also associated with recurrence, possibly due to exposure to broad-spectrum antibiotics^38^. While sepsis has been less studied in a recurrence context, prior infections have been highlighted as a key risk factor for primary CDI^3^.

Our findings suggest that disease onset and association continues to matter to recurrence and mortality risks for CDI patients, but in a way that has changed over time, with recurrence risks increasing for patients with COCA disease compared to HOHA disease. Previous studies have found inconsistent associations between initial CDI acquisition site and recurrence^16,17^. While a recent systematic review and meta-analysis using global data 2016-2024 found higher recurrence rates for community-associated CDI^39^, in keeping with later data from our study, another systematic review (based on studies published until 2019) found consistently higher recurrence rates for healthcare-associated CDI (analogous to HOHA+COHA in our study) across countries^1^. Nevertheless, neither of these studies formally addressed the competing risk of death, which our study does, as well as spanning the full time period, potentially explaining the previous apparently conflicting observations.

Although power was limited, our more detailed investigation suggests that the shift in recurrence risks occurred before COVID-19, appearing to coincide with decline of the highly virulent ribotype-027 strain which was particularly associated with hospital-associated CDI. This might attenuate differences between recurrence risks for community and hospital associated CDI, as the causative strains became more similar. It is therefore surprising that we found a greater risk of recurrence after community-associated infection, for unclear reasons despite several potential explanations.

First, testing policy may have changed over time, contributing to increased ascertainment of previously missed COCA index cases and recurrences. Risk factors for community-associated CDI are generally less well-understood but include medications such as proton-pump inhibitors and antibiotics, living near farms or livestock, and specific conditions including kidney disease^40-42^.

Increased recurrence risks for COCA cases could therefore relate to changes in community prescriptions. Other potential drivers include differences in treatment between hospital and community settings, including antibiotic choices such as fidaxomicin (which reduces recurrence risk^43^), likely less commonly used in primary care. Our results suggest that elevated COCA vs HOHA recurrence risks persist when controlling for admission within 48h of sample collection. Nevertheless, initial treatment may still matter and patients tested in the community might go longer before receiving a CDI diagnosis. Delayed treatment has been associated with progression to severe and fulminant CDI^44^, and longer gaps between symptom onset and CDI testing are common in the community compared to hospitals^45^. Furthermore, continuing non-CDI antibiotics after the index positive test is strongly associated with recurrence^16,19,46^, which could be more likely in the community, where patients are less closely monitored.

In our study, community hospital cases could not explain the increased COCA recurrence risk: these admissions are not counted as inpatients and therefore do not contribute to surveillance classifications despite substantial exposure to healthcare and its associated risks. Over time, we instead observed an increase in the proportion of community-onset samples from primary care. Nevertheless, COCA cases may increasingly be receiving other forms of risk-carrying healthcare exposure which is not captured, including from nursing homes. Given increased competition for hospital beds, some patients who are now classified as COCA might have had recent hospitalisations in earlier periods with less bed pressures. That is, the difference between COCA and non-COCA patients may have decreased over time due to strain on the healthcare system.

Baseline blood test results were not associated with recurrence in our study, but, as expected, we observed several associations with death before recurrence^47,48^. Index severity may matter more to mortality than recurrence risk, and/or other variables may better capture infection severity, including vital signs. In our study, these were not available before 2016, so not included. We did not include antibiotic or other medication exposures for the same reasons. While antibiotics, including prescribed in the community^49,50^, remain a crucial risk factor for CDI, instead we were able to study a long timeframe of nearly 25 years. Due to substantial missingness, we also were unable to include body mass index (also only available after 2016) and ethnicity in our models. Furthermore, while our study failed to detect significant changes in risk factors other than surveillance classification over time, this might be due to limited power rather than a lack of real heterogeneity.

Another key limitation is that this study was conducted in one region, covering ∼1% of the UK. Hospital exposures and CDI recurrence were only ascertained using OUH data, so patients with admissions or positive CDI tests outside the region would not be captured. We adjusted for ascertainment bias by restricting to catchment area, but note that recurrence would likely be underestimated more in the community than in hospital settings. Ascertainment of medical conditions was restricted to diagnostic codes from consultant episodes ending before sample collection to ensure conditions were present before sampling: this could lead to underestimation, especially of acute conditions. Large changes to coding practice, particularly between 2000-2012 will also have improved comorbidity ascertainment, potentially influencing estimates. Any conditions developing post-index case were not included: a time-updated model including these and other time-varying factors such as treatment response might explain more recurrence risk but would not allow baseline interventions to be targeted.

In conclusion, this study has highlighted a shift towards increased recurrence risks for patients with community-associated CDI compared to hospital-onset hospital-associated CDI. This change in trend appears to have begun before COVID-19, and other risk factors for recurrence appeared relatively stable over time. Recurrence remains a major challenge for COCA CDI patients and further research is needed to understand the best avenues for prevention.

## Acknowledgements

This work uses data provided by patients and collected by the UK’s National Health Service as part of their care and support. We thank all the people of Oxfordshire who contribute to the Infections in Oxfordshire Research Database. We also thank the Patient and Public Involvement (PPI) panel for this NIHR Doctoral Fellowship for their input.

## Author contributions

The specific analysis was originally designed by ASW, DWE, and ED, with ongoing input from all authors on analysis design and results. ED conducted the statistical analysis, and ED, ASW and DWE drafted the manuscript. All authors contributed to interpretation of the data and results, manuscript revision, and approved the final version of the manuscript.

## Data Availability

The datasets analysed in this study are not publicly available as they contain personal data, but are available from the Infections in Oxfordshire Research Database (IORD | NIHR Biomedical Research Centre: Oxford), subject to an application and research proposal meeting the ethical and governance requirements of the Database. For further details on how to apply for data access and for a research proposal template please.

