## Supplementary tables and figures for "Increasing risks of recurrence following community-associated *Clostridioides difficile* infection in Oxfordshire 2000-2025"

**Supplementary Table S1: Further characteristics of study periods**

| **Period name** | **Late entry criteria** | **Late entries*** | **Patients with missing covariates  (tests dropped)** | **Recurrences in period** | **Deaths in period** | **Censored in period** | **Days to recurrence in recurrences** | **Days to death before recurrence in deaths before recurrence** | **Days to censoring in those censored** | **Patients with ≥1 recurrence** |
| --- | --- | --- | --- | --- | --- | --- | --- | --- | --- | --- |
| *Full study* | - | - | 285 (297) | 1,410 (14.9%) | 6,375 (67.4%) | 1,671 (17.7%) | 57  (38, 130) | 143  (21, 1,053) | 4,982  (1,952, 6,596) | 1,116 (13.9%) |
| *High virulence* | ELISA-positive toxin test between 01 April 2002 and 31 March 2003 and still alive by 01 April 2003 | 360 | 41 (41) | 587 (13.6%) | 2,228 (51.5%) | 1,509 (34.9%) | 52 (37, 102) | 35 (10, 215) | 800  (298, 1,375) | 465 (12.4%) |
| *Post-high virulence* | ELISA-positive toxin test between 01 April 2007 and 31 March 2008 and still alive by 01 April 2008 | 430 | 34 (34) | 267 (12.9%) | 861 (41.7%) | 937 (45.4%) | 53 (37, 97) | 80 (19, 353) | 931  (536, 1,461) | 215 (12.0%) |
| *Pre-pandemic* | ELISA-positive toxin test between 01 April 2011 and 31 March 2012 and still alive by 01 April 2012 | 166 | 14 (15) | 155 (11.5%) | 620 (45.9%) | 576  (42.6%) | 56 (36, 90) | 110 (25, 599) | 1,641 (710, 2,645) | 121 (10.1%) |
| *Post-Pandemic* | GDH and toxin EIA-positive test between 01 March 2019 and 29 February 2020 and still alive by June 01 2020 | 70 | 5 (5) | 117  (13.3%) | 346  (39.3%) | 418 (47.4%) | 56 (36, 89) | 78 (26, 302) | 661  (259, 1,336) | 94 (12.3%) |

* positive test before the start of the current period; contributed follow-up time to recurrence/death within this period

Note: showing N (%) or median (IQR). Days to death before recurrence in deaths before recurrence affected by longer follow-up periods given age of CDI cases.

**Supplementary Table S2: Other characteristics by study period**

| **Factor** | *Overall*  *2000-2025* | *“High virulence”*  *2003-2008* | *“Post high virulence”*  *2008-2012* | *“Pre-pandemic”*  *2012-2020* | *“Post-pandemic”*  *2020-2025* |
| --- | --- | --- | --- | --- | --- |
| Elixhauser comorbidity index (median, IQR) | 5 (0, 13) | 4 (0, 11) | 5 (0, 12) | 7 (0, 17) | 10 (0, 21) |
| Elective bed-days (weeks in past 12 months) (median, IQR) | 0.00 (0.00, 0.05) | 0.00 (0.00, 0.07) | 0.00 (0.00, 0.06) | 0.00 (0.00, 0.06) | 0.00 (0.00, 0.03) |
| Recent surgical admission (past 3 months) (n, %) | 1,571 (16.6%) | 717 (16.6%) | 365 (17.7%) | 257 (19.0%) | 158 (17.9%) |
| Recent GI admission (past 3 months) (n, %) | 485 (5.1%) | 232 (5.4%) | 153 (7.4%) | 88 (6.5%) | 45 (5.1%) |
| Palliative care (past 5 years) (n, %) | 183 (1.9%) | 19 (0.4%) | 27 (1.3%) | 53 (3.9%) | 77 (8.7%) |
| Cancer (past 5 years) (n, %) | 1,920 (20.3%) | 734 (17.0%) | 464 (22.5%) | 393 (29.1%) | 274 (31.1%) |
| Dementia (past 5 years) (n, %) | 1,003 (10.6%) | 409 (9.5%) | 236 (11.4%) | 183 (13.5%) | 127 (14.4%) |
| Anaemia (past 5 years) (n, %) | 1,766 (18.7%) | 620 (14.3%) | 403 (19.5%) | 354 (26.2%) | 335 (38.0%) |
| Heart failure (past 5 years) (n, %) | 1,220 (12.9%) | 481 (11.1%) | 224 (10.8%) | 225 (16.7%) | 176 (20.0%) |
| Hypertension (past 5 years) (n, %) | 3,132 (33.1%) | 1,196 (27.7%) | 829 (40.1%) | 696 (51.5%) | 488 (55.4%) |
| Other heart disease (past 5 years) (n, %) | 3,599 (38.1%) | 1,379 (31.9%) | 861 (41.7%) | 700 (51.8%) | 508 (57.7%) |
| COPD (past 5 years) (n, %) | 999 (10.6%) | 403 (9.3%) | 185 (9.0%) | 201 (14.9%) | 138 (15.7%) |
| Oesophageal disease (past 5 years) (n, %) | 857 (9.1%) | 330 (7.6%) | 130 (6.3%) | 196 (14.5%) | 188 (21.3%) |
| Liver disease (past 5 years) (n, %) | 1,178 (12.5%) | 397 (9.2%) | 277 (13.4%) | 251 (18.6%) | 264 (30.0%) |
| Cerebrovascular disease (past 5 years) (n, %) | 477 (5.0%) | 151 (3.5%) | 105 (5.1%) | 93 (6.9%) | 121 (13.7%) |
| Diabetes (past 5 years) (n, %) | 1,453 (15.4%) | 614 (14.2%) | 328 (15.9%) | 275 (20.4%) | 204 (23.2%) |
| Acute renal failure (past 3 months) (n, %) | 851 (9.0%) | 252 (5.8%) | 139 (6.7%) | 231 (17.1%) | 202 (22.9%) |
| Pneumonia/bronchitis (past 3 months) (n, %) | 1,466 (15.5%) | 655 (15.1%) | 289 (14.0%) | 267 (19.8%) | 172 (19.5%) |
| UTI (past 3 months) (n, %) | 1,100 (11.6%) | 434 (10.0%) | 230 (11.1%) | 226 (16.7%) | 162 (18.4%) |
| Transplant (past 5 years) (n, %) | 216 (2.3%) | 80 (1.9%) | 68 (3.3%) | 40 (3.0%) | 42 (4.8%) |
| Missing FBC (n, %) | 3,113 (32.9%) | 1,429 (33.0%) | 707 (34.2%) | 425 (31.5%) | 230 (26.1%) |
| Missing Creatinine (n, %) | 2,931 (31.0%) | 1,305 (30.2%) | 691 (33.5%) | 413 (30.6%) | 222 (25.2%) |
| Missing CRP (n, %) | 3,421 (36.2%) | 1,535 (35.5%) | 807 (39.1%) | 465 (34.4%) | 242 (27.5%) |
| Missing Albumin (n, %) | 3,912 (41.4%) | 1,840 (42.6%) | 902 (43.7%) | 540 (40.0%) | 277 (31.4%) |
| Eosinophils (x10^9^/L) (median, IQR) | 0.09 (0, 0.19) | 0.08 (0, 10.19) | 0.09 (0, 0.19) | 0.08 (0, 0.20) | 0.08 (0.03, 0.17) |
| Lymphocytes (x10^9^/L) (median, IQR) | 1.12 (0.75, 1.60) | 1.13 (0.79, 1.61) | 1.18 (0.82, 1.70) | 1.14 (0.77, 1.68) | 1.04 (0.73, 1.55) |
| Haemoglobin (x10^9^/L) (median, IQR) | 109 (97, 123) | 110 (98, 124) | 109 (97, 122) | 109 (95, 123) | 107 (91, 120) |
| Platelets (x10^9^/L) (median, IQR) | 300 (211, 406) | 307 (216, 409) | 296 (206, 393) | 290 (203, 388) | 272 (189, 376) |
| Creatinine (umol/L) (median, IQR) | 91 (68, 141) | 92 (71, 145) | 89 (68, 126) | 86 (62, 130) | 78 (56, 126) |
| Albumin (g/L) median(IQR) | 25 (20.0, 29.2) | 24.1 (20.0, 28.2) | 26.2 (22.1, 30.2) | 25.0 (21.0, 29.2) | 26.0 (21.0, 30.0) |

**Supplementary Table S3: Results from variable selection on Fine-Gray models of recurrence and death before recurrence in four different study periods**

| **Factor** | | **“Post-pandemic”** | | **“Pre-pandemic”** | | **“Late ELISA”** | | **“High virulence”** | |
| --- | --- | --- | --- | --- | --- | --- | --- | --- | --- |
|  |  | Recurrence | Death before recurrence | Recurrence | Death  before recurrence | Recurrence | Death  before recurrence | Recurrence | Death  before recurrence |
| Surveillance classification | | x | x |  | x | x | x |  | x |
| Demographics | Age |  | x |  | x | x | x | x | x |
|  | Sex |  |  |  | x |  | x |  | x |
|  | Deprivation |  | x | x |  |  | x |  |  |
|  | Catchment |  |  | x |  | x | x | x |  |
| Prior test history  (past 6 months) | Prior CDI |  |  |  |  |  |  | x | x |
|  | Prior negative test |  |  | x |  |  |  |  |  |
|  | Prior carriage |  |  |  | x |  |  |  |  |
| Comorbidity scores | Charlson |  |  |  | x | x | x |  | x |
|  | Elixhauser |  | x | x | x |  | x | x | x |
|  | Gilbert (Frailty) | x |  |  | x |  |  | x |  |
| Hospital exposures | Emergency bed-days  (past 12 months) | x |  | x |  | x |  | x | x |
|  | Elective bed-days  (past 12 months) |  |  |  |  |  |  |  | x |
|  | Number of emergency admissions  (past 6 months) | x |  |  |  |  | x |  |  |
|  | Number of outpatient appointments  (past 6 months) | x | x | x |  | x |  | x |  |
|  | Recent surgical admission  (past 3 months) |  |  |  |  |  | x |  | x |
| Chronic conditions  (past 5 years) | Palliative care |  | x | x | x |  | x |  |  |
|  | Immunosuppression |  | x | x | x | x |  |  | x |
|  | Cancer |  |  |  |  |  |  |  | x |
|  | Dementia |  | x |  |  |  |  | x |  |
|  | Anaemia |  |  |  | x |  | x |  |  |
|  | Chronic kidney disease | x |  | x |  | x |  | x |  |
|  | Heart failure |  |  |  | x |  | x |  |  |
|  | Hypertension |  |  |  |  | x |  |  |  |
|  | Other heart disease | x |  |  | x |  |  |  |  |
|  | COPD |  | x |  |  |  |  |  |  |
|  | Inflammatory bowel disease | x |  |  |  |  |  |  |  |
|  | Diverticular disease |  | x |  |  |  | x | x |  |
|  | Oesophageal disease |  |  |  | x |  |  |  | x |
| Acute conditions  (past 3 months) | Acute renal failure |  | x |  |  |  |  |  |  |
|  | Pneumonia/bronchitis |  | x |  | x |  | x |  |  |
|  | Sepsis/bacteraemia | x |  |  | x |  |  | x |  |
|  | UTI |  |  |  |  | x |  |  |  |
| Procedure history  (past 5 years) | Transplant | x |  |  | x |  |  |  |  |
|  | Feeding tubes |  | x |  |  |  |  | x |  |

Note: Variable selection was performed with a threshold of p<0.1 individually on each of the eight models. Variables evaluated but not selected by any models were: cerebrovascular disease (diagnosis in past 5 years), liver disease (diagnosis in past 5 years), diabetes (diagnosis in past 5 years), and recent GI admission (past 3 months). Variables only selected by a single model (marked in red) were excluded from further modelling.

**Supplementary Table S4. Patient location at time of sample collection for CDI by study period**

|  | **Hospital onset (HOHA)** | **Community onset (COHA, COIA, COCA)** | | | |
| --- | --- | --- | --- | --- | --- |
| **Period** | **Hospital** | **Hospital** | **Community hospital** | **Primary care** | **Unknown** |
| *Overall study 2000 - 2025* | 5,644 (59.7%) | 1,932 (20.4%) | 612 (6.5%) | 1,263 (13.4%) | 5 (0.1%) |
| *High virulence 2003 – 2008* | 2,893 (66.9%) | 702 (16.2%) | 339 (7.8%) | 387 (9.0%) | 3 (0.1%) |
| *Post-high virulence 2008 – 2012* | 1,035 (50.1%) | 434 (21.0%) | 138 (6.7%) | 458 (22.2%) | 0 (0.0%) |
| *Pre-pandemic 2012 - 2020* | 550 (40.7%) | 397 (29.4%) | 63 (4.7%) | 341 (25.2%) | 0 (0.0%) |
| *Post-pandemic 2020 – 2025* | 380 (43.1%) | 273 (31.0%) | 47 (5.3%) | 181 (20.5%) | 0 (0.0%) |

NOTE: Trends by year are shown in Figure S15. “Hospital” includes OUH hospitals, private hospitals, and nearby non-OUH hospitals, “community hospital” includes community hospitals and hospice, “primary care” includes primary care practitioners (GPs), nursing homes, and unknown non-hospital locations.

**Supplementary Figure S1. De-duplicated CDI test results in adults performed at OUH NHS Trust April 2000 to March 2025**

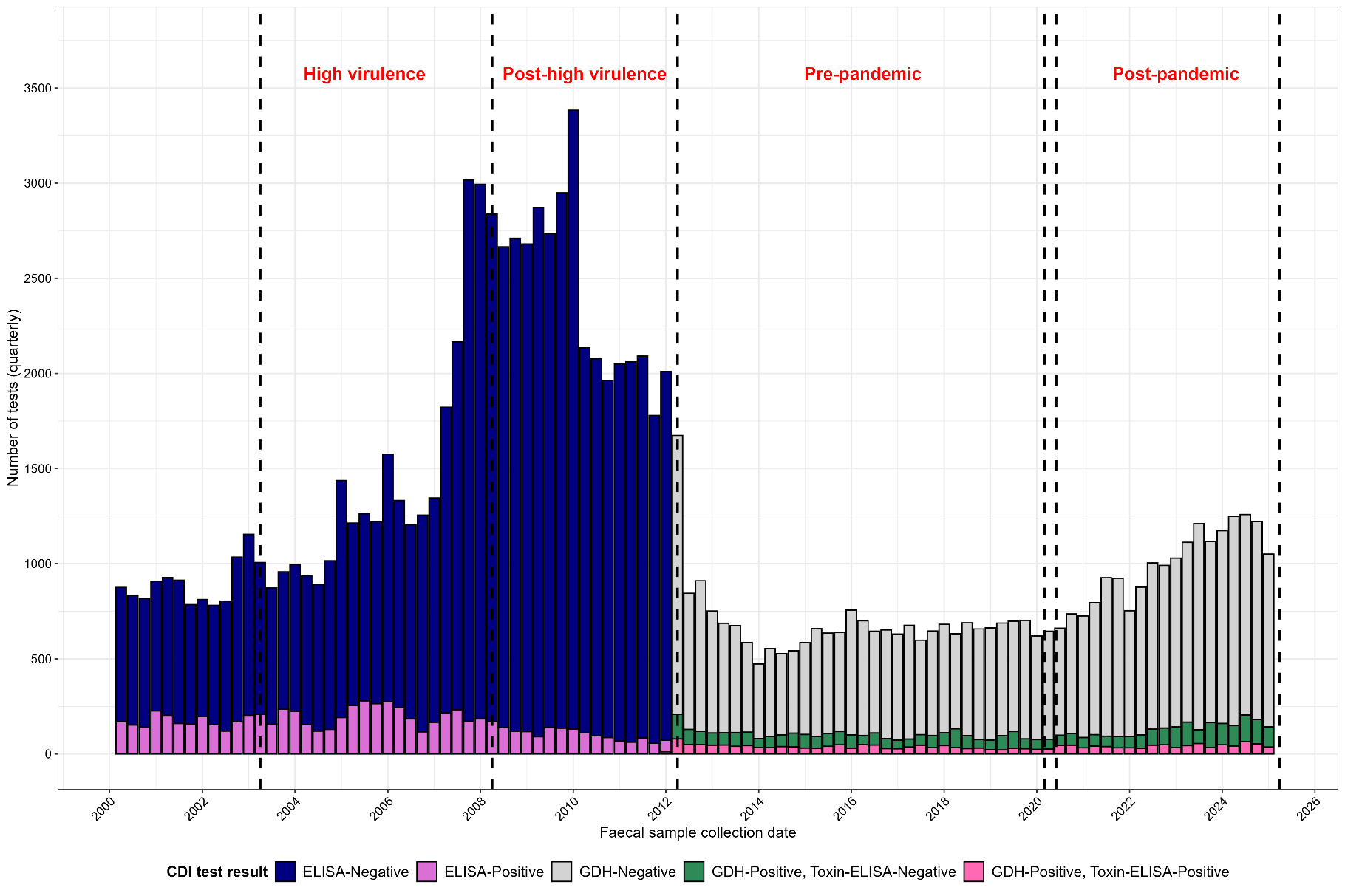

Note: All tests de-duplicated to 28 days from prior CDI. Negative tests and GDH-positive toxin-negative (carriage) not de-duplicated to previous negative or carriage. Substantial numbers of negative tests in the “post-high virulence” period due to repeat testing and concern about sensitivity of the standalone ELISA test.

**Supplementary Figure S2. Cumulative 12-month count of adult CDI cases April 2001 – March 2025**

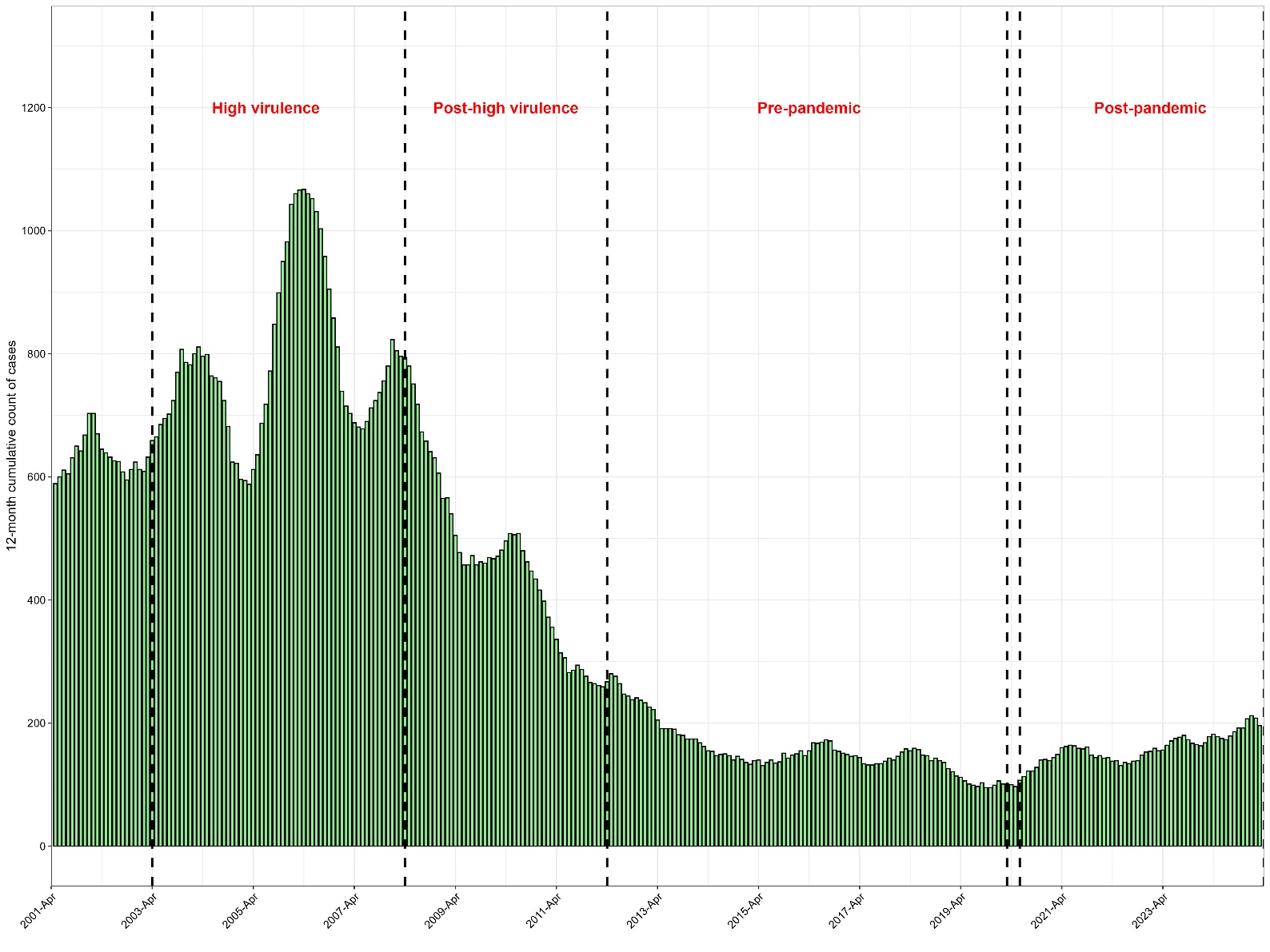

NOTE: A small number of tests from adult patients with missing address or sex were excluded.

**Supplementary Figure S3. Cumulative 12-month count of CDI recurrences, April 2001 – March 2025**

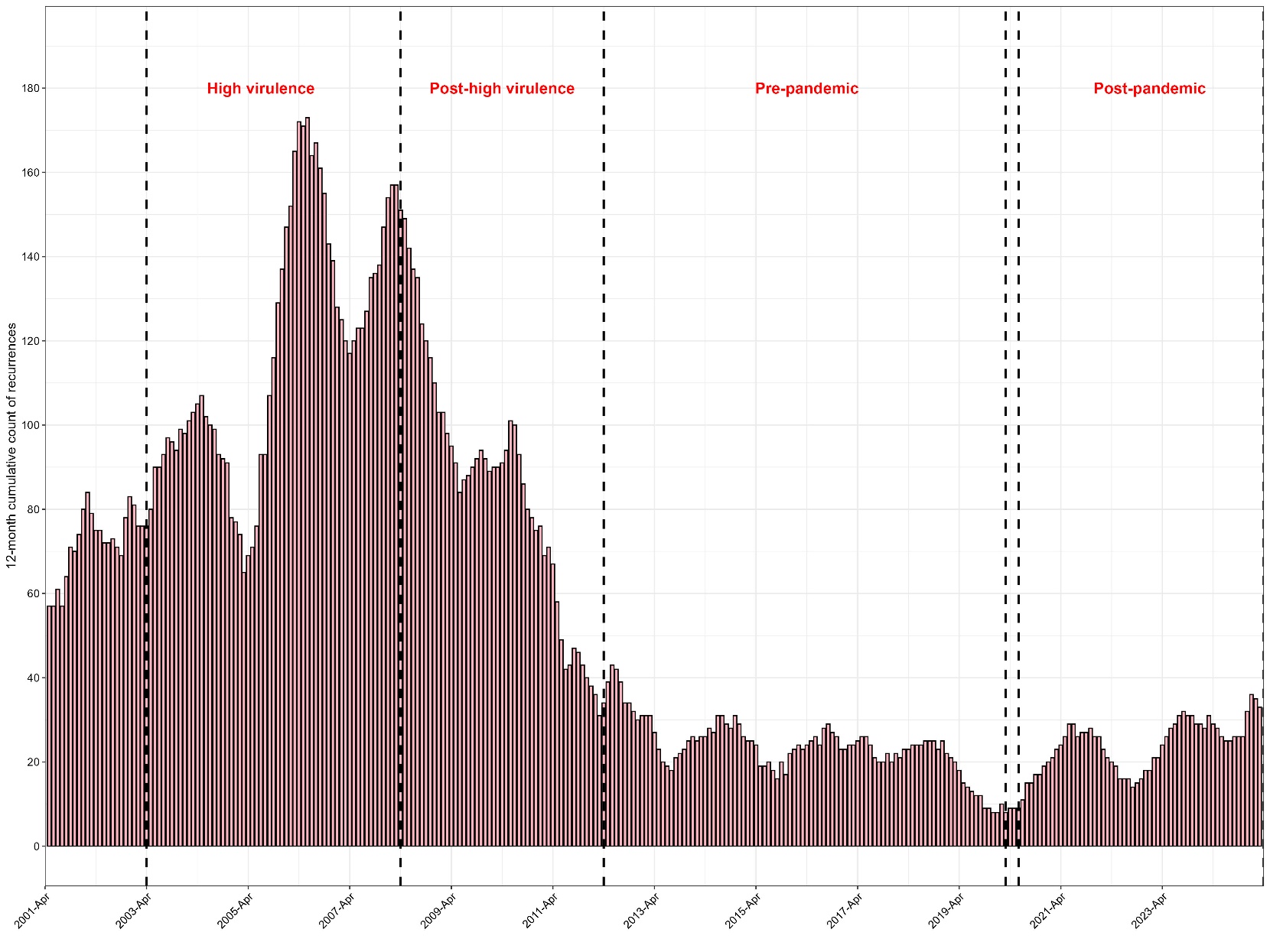

NOTE: A small number of tests from adult patients with missing address or sex were excluded.

**Supplementary Figure S4: Cumulative incidence of recurrence and death before recurrence over the first 3 years post-episode**

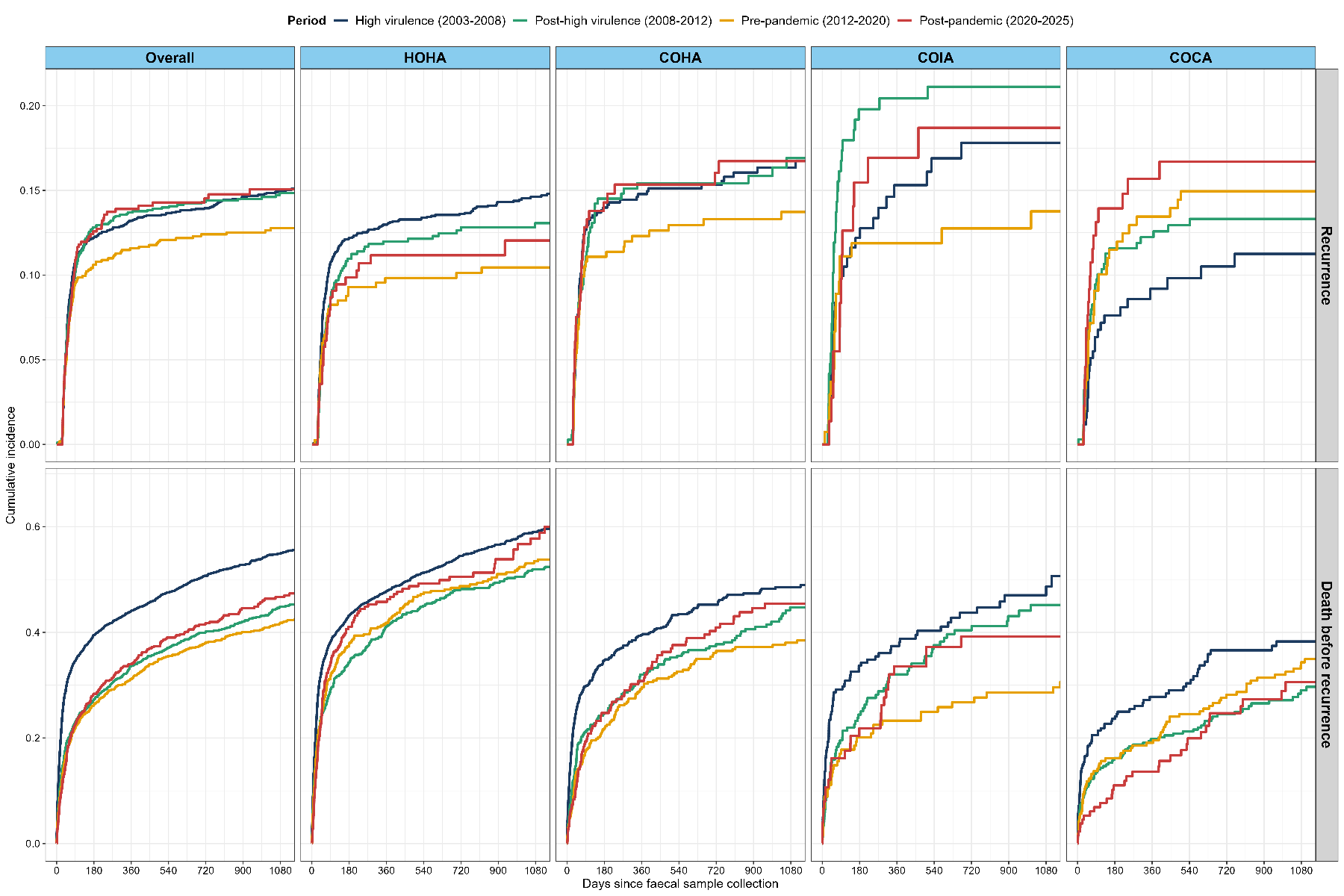

NOTE: Follow-up truncated at 3 years. Estimates by Aalen-Johansen with the other outcome as a competing risk, analysis restricted to patients with OUH catchment ≥80% to minimise ascertainment bias

**Supplementary Figure S6: Risk factors for recurrence and death before recurrence before variable selection on period-interactions**

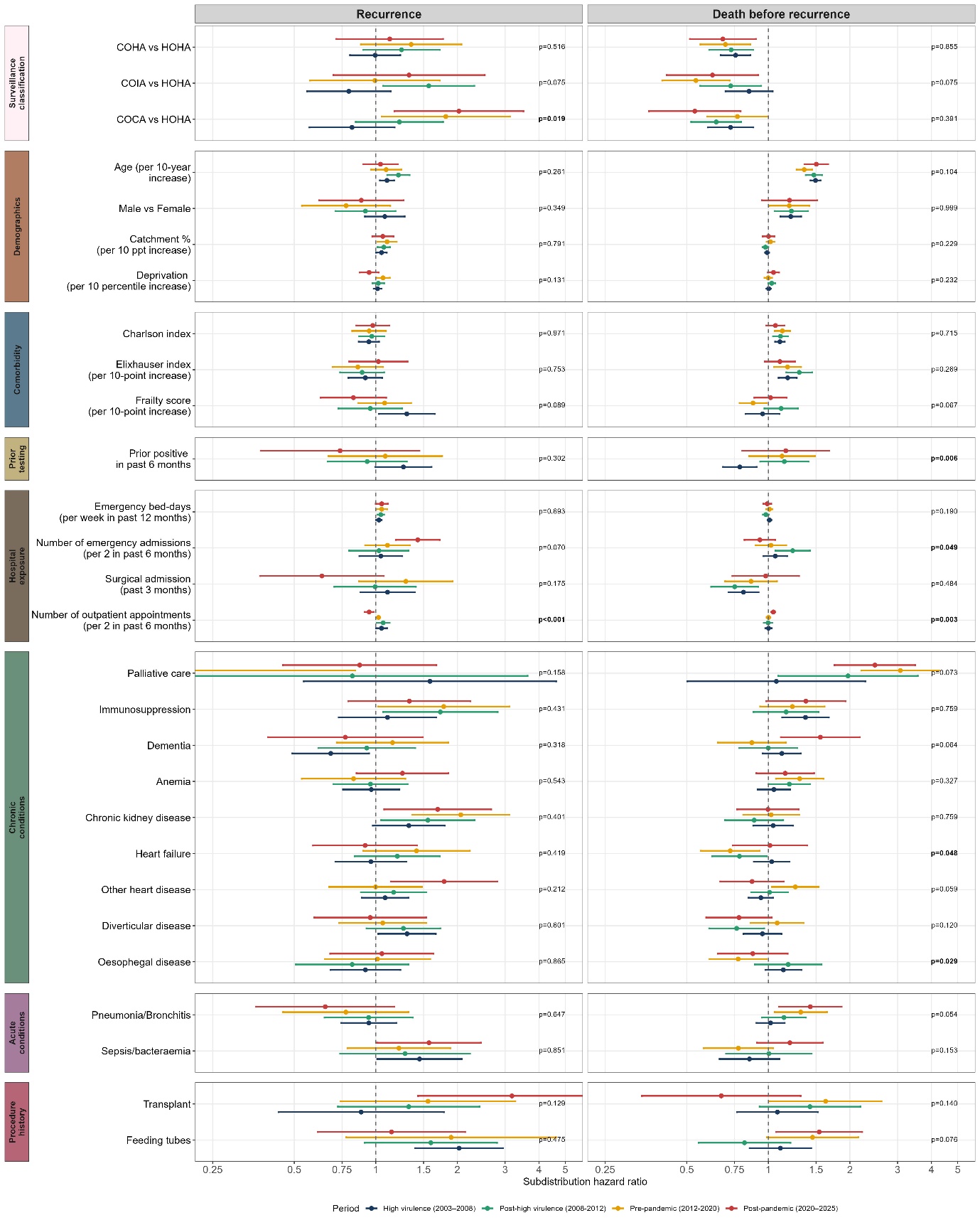

Note: All variables selected by backward selection in two or more models (Table S3) included with period-specific effects (heterogeneity p-values testing for variation in the effect of each factor across the 4 study periods shown on the right side of each plot). Overall heterogeneity p-values for surveillance classification were 0.048 (recurrence) and 0.319 (death). Plot truncated at (0.25, 0.5). The point estimate and lower 95% CI for palliative care in “pre-pandemic” (recurrence) were 0.13 and 0.02 respectively. The lower 95% CI for palliative care in “late ELISA” (recurrence) was 0.19. The upper 95% CI for transplant in “post-pandemic” (recurrence) was 7.06. Robust standard errors were clustered by patient.

**Supplementary Figure S7: Risk factors for recurrence and death before recurrence before variable selection on main effects**

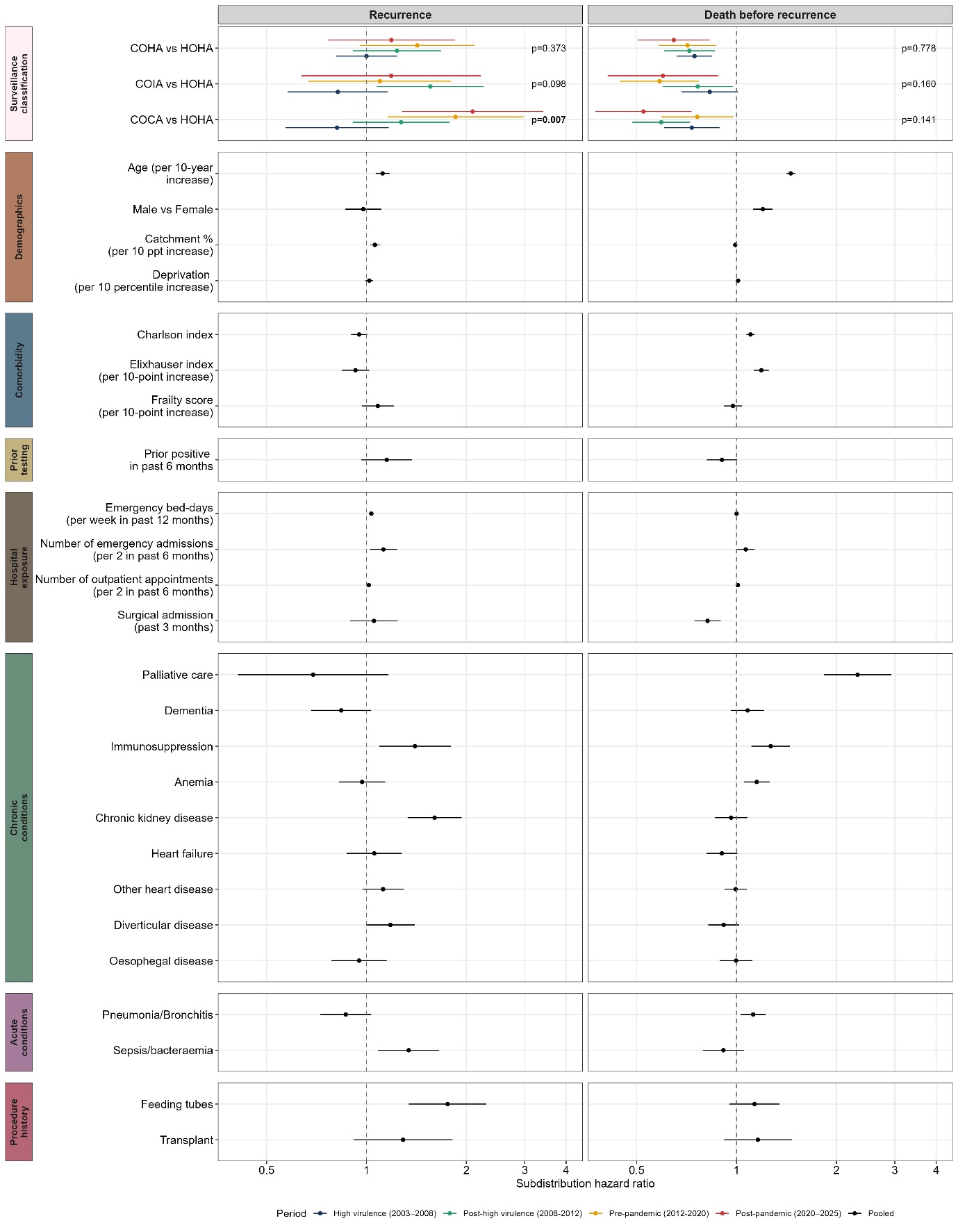

Note: Overall heterogeneity p-values for surveillance classification were 0.021 (recurrence) and 0.250 (death). Models were stratified by period, and robust standard errors were clustered by patient. For comparability, fitting the same model as identified for recurrence to death before recurrence, to assess the potential for competing risks.

**Supplementary Figure S8. Cause-specific risk factors for recurrence and death before recurrence**

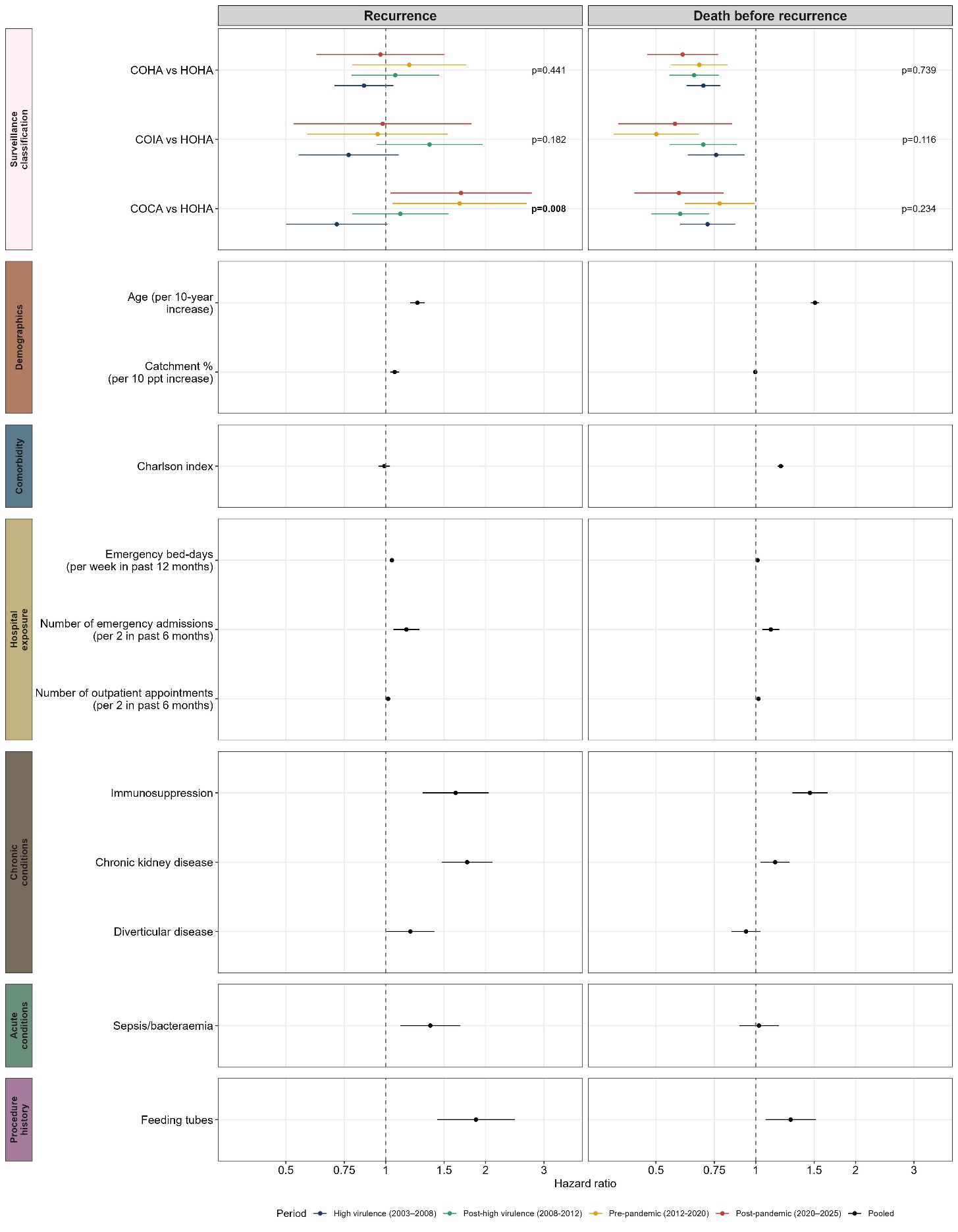

Note: Overall heterogeneity p-values for surveillance classification were 0.036 (recurrence) and 0.220 (death). Models were stratified by period, and robust standard errors were clustered by patient.

**Supplementary Figure S9. Surveillance classification of initial and subsequent CDI episodes and final outcome for the full study period**

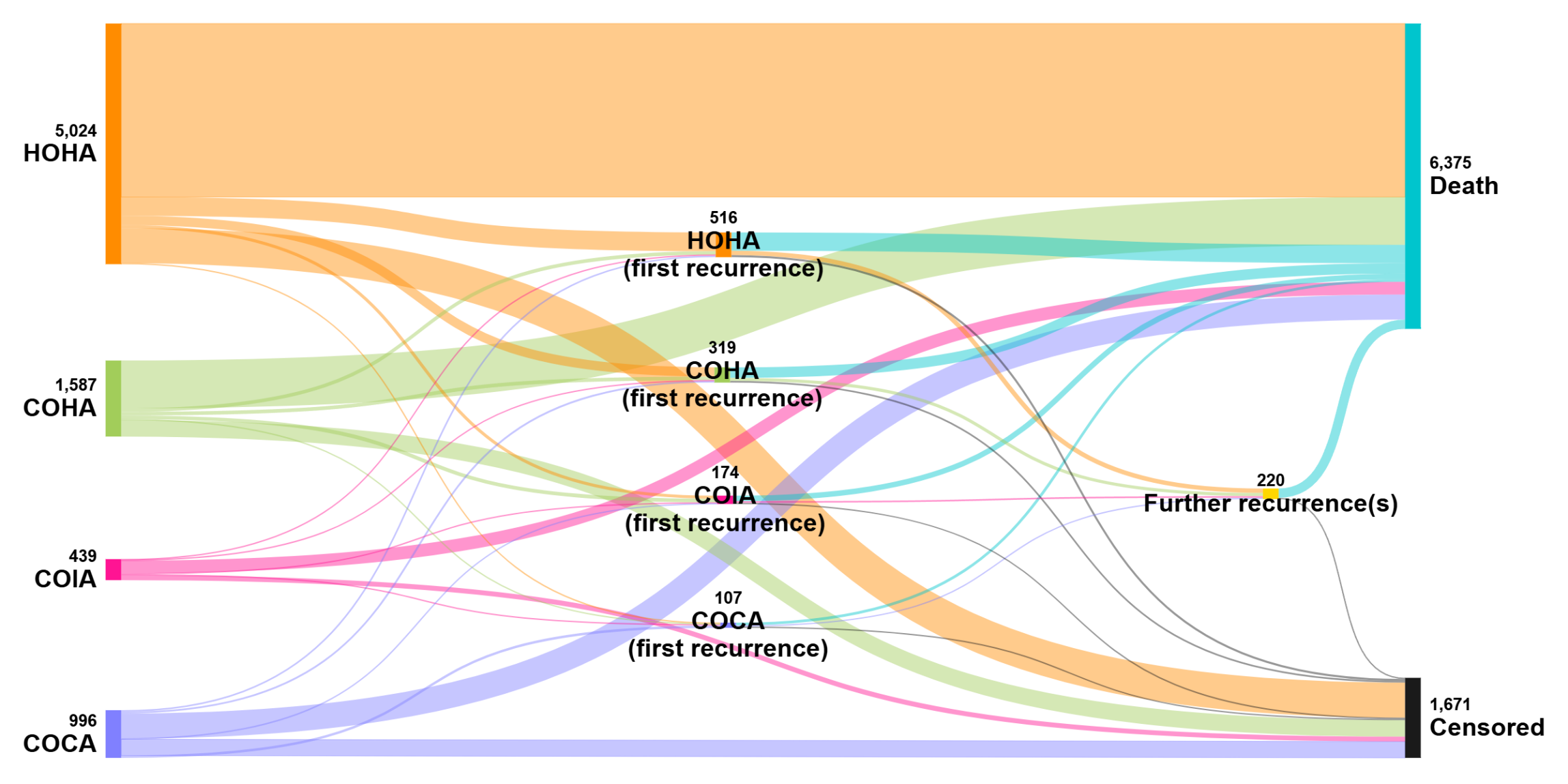

**Supplementary Figure S10: Recurrence risk by surveillance classification over continuous calendar time**

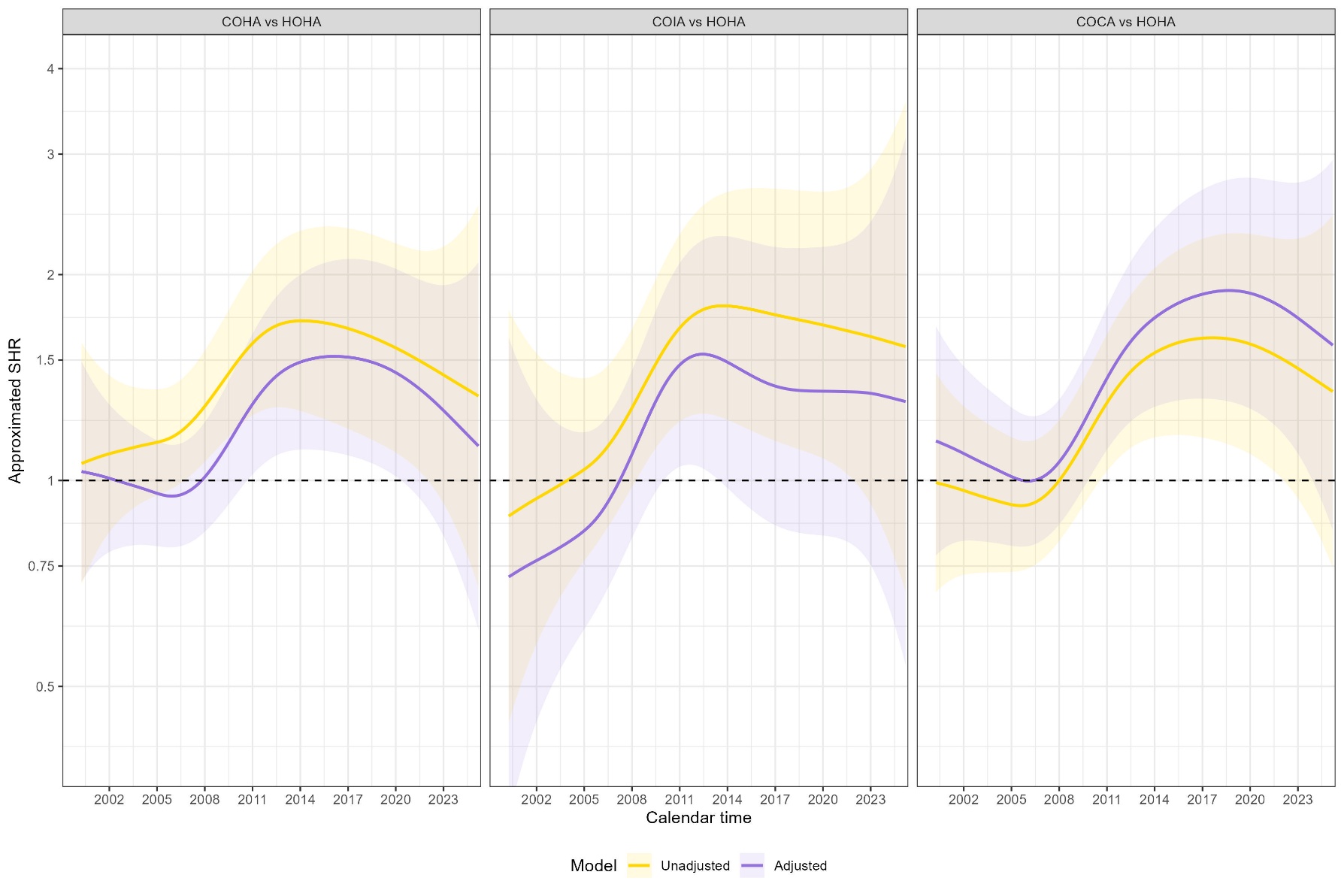

NOTE: Estimates for fixed effects of other covariates in the adjusted model (purple) are given in Figure S11. Models are fit as GAMs on modified survival data that allows for approximation of subdistribution hazards using a cause-specific Cox model specification

**Supplementary Figure S11: Fixed effects from adjusted continuous time model displayed in Figure S10**

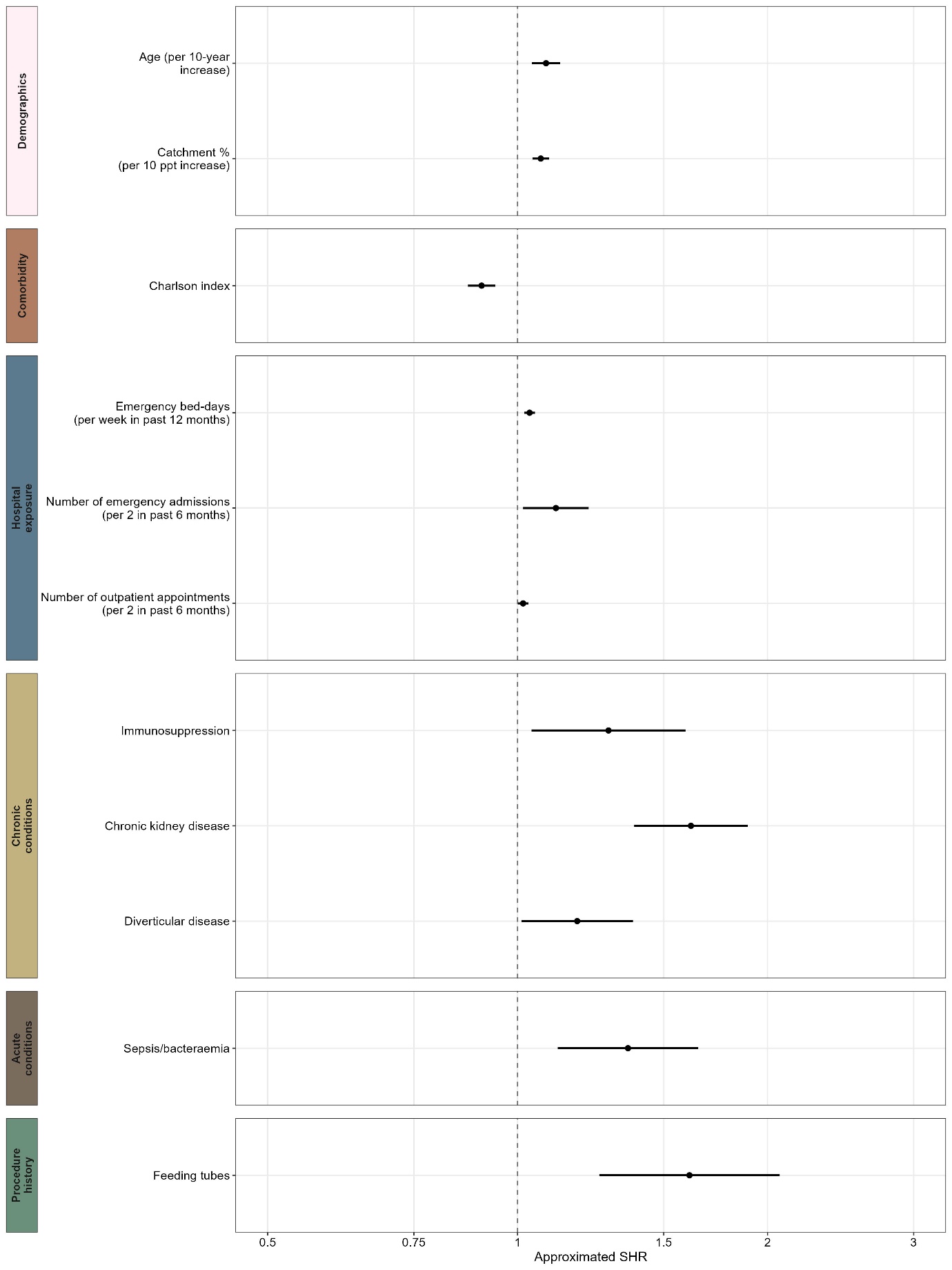

**Supplementary Figure S12. Risk factors for recurrence and death before recurrence, follow-up censored at 365 days**

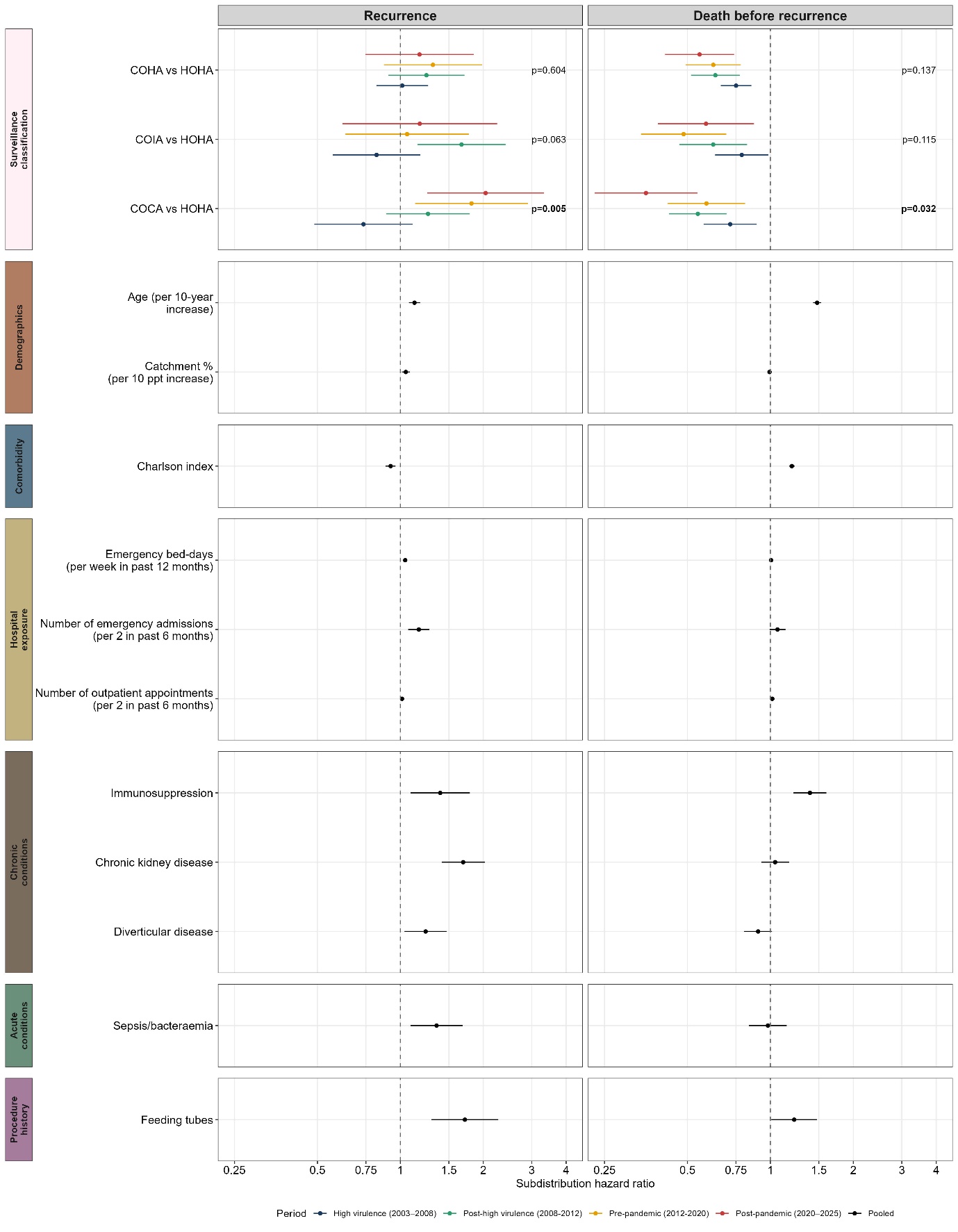

Note: Overall heterogeneity p-values for surveillance classification were 0.014 (recurrence) and 0.053 (death). Models were stratified by period, and robust standard errors were clustered by patient.

**Supplementary Figure S13. Risk factors for recurrence and death before recurrence, follow-up censored at 90 days**

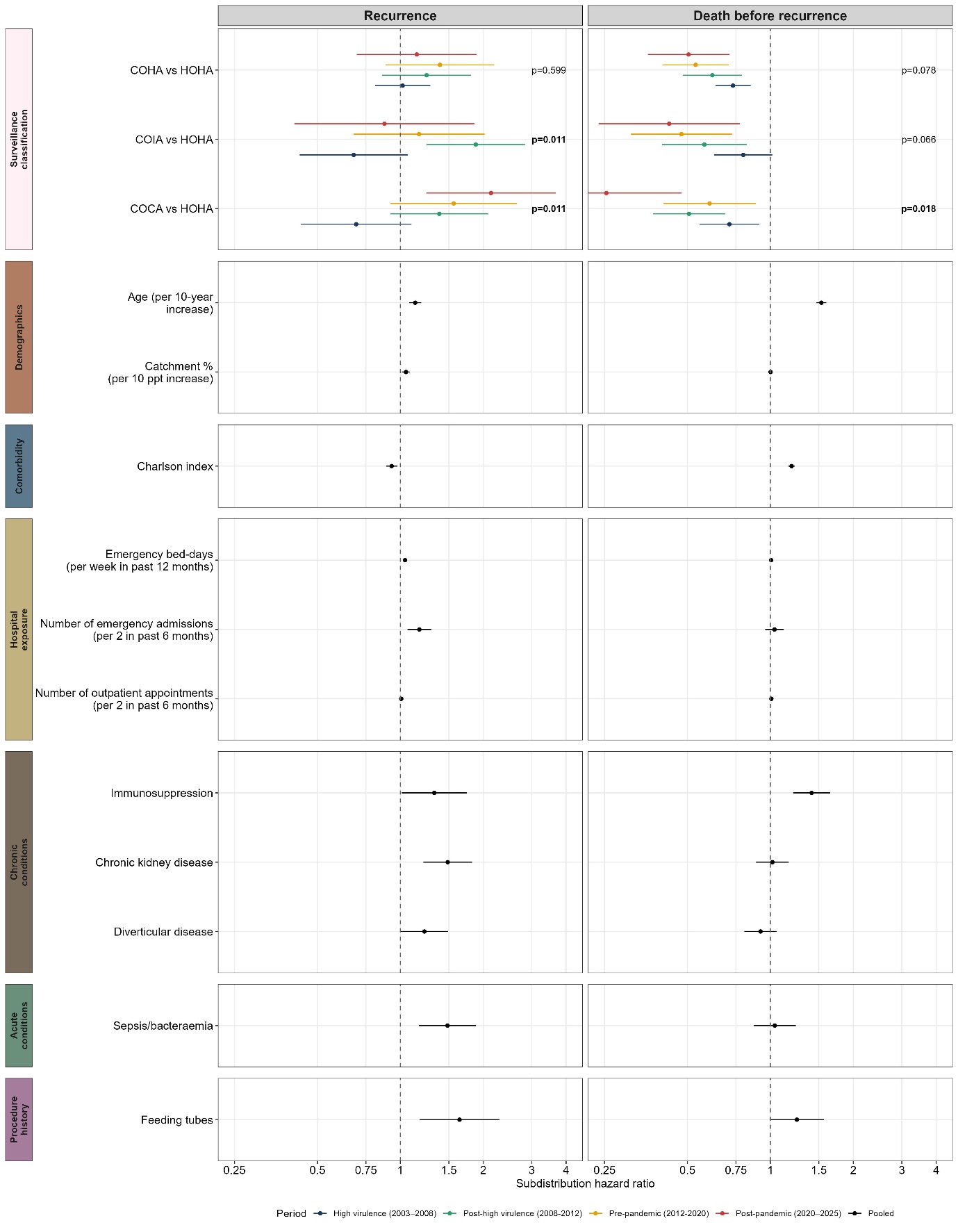

Note: Overall heterogeneity p-values for surveillance classification were 0.006 (recurrence) and 0.017 (death). Models were stratified by period, and robust standard errors were clustered by patient. The lower 95% CI for COCA vs HOHA in the “post-pandemic” period is 0.14 (death).

**Supplementary Figure S14. Cause-specific risk factors for recurrence and death before recurrence, follow-up censored at 365 days**

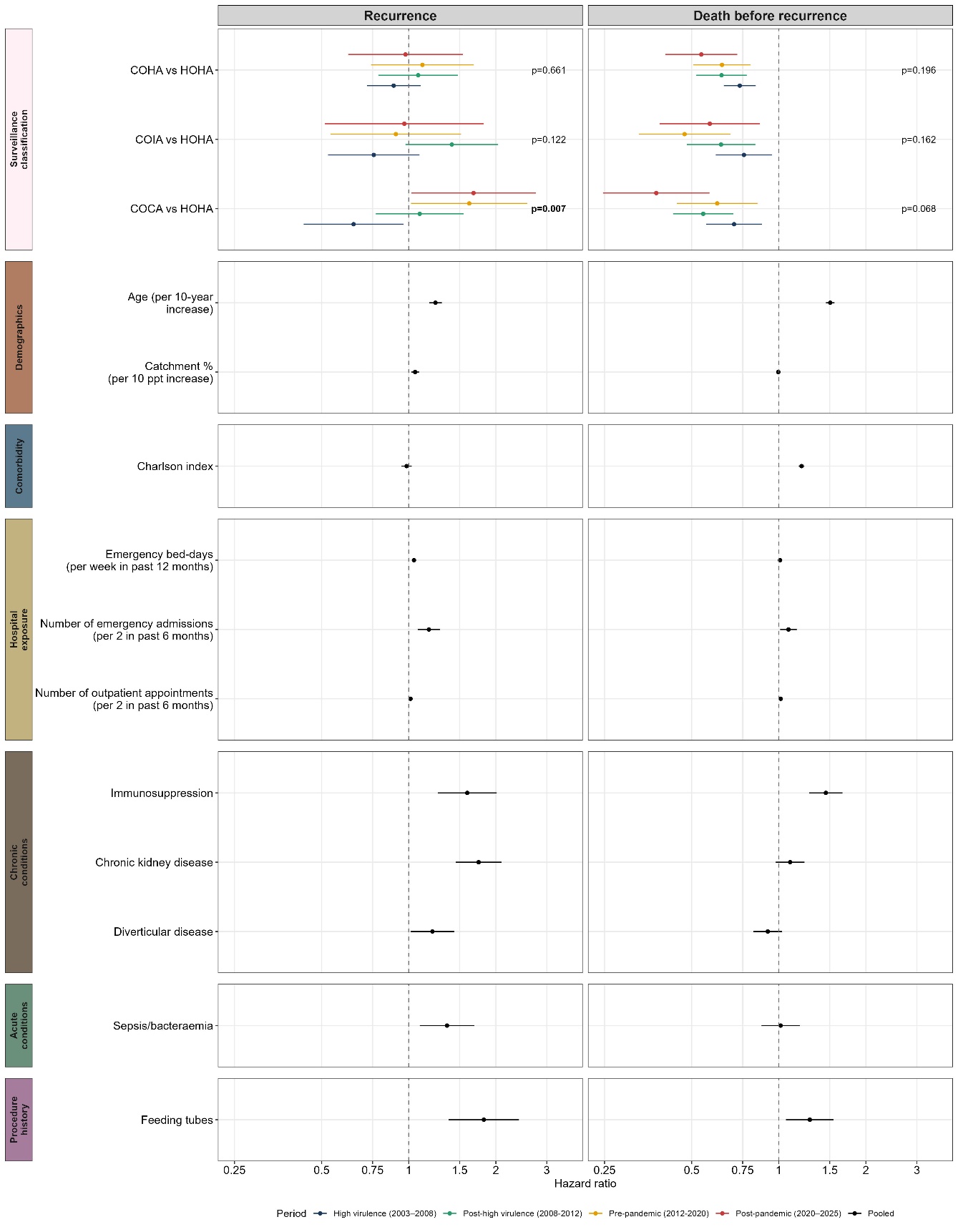

Note: Overall heterogeneity p-values for surveillance classification were 0.024 (recurrence) and 0.112 (death). Models were stratified by period, and robust standard errors were clustered by patient.

**Supplementary Figure S15. Cause-specific risk factors for recurrence and death before recurrence, follow-up censored at 90 days**

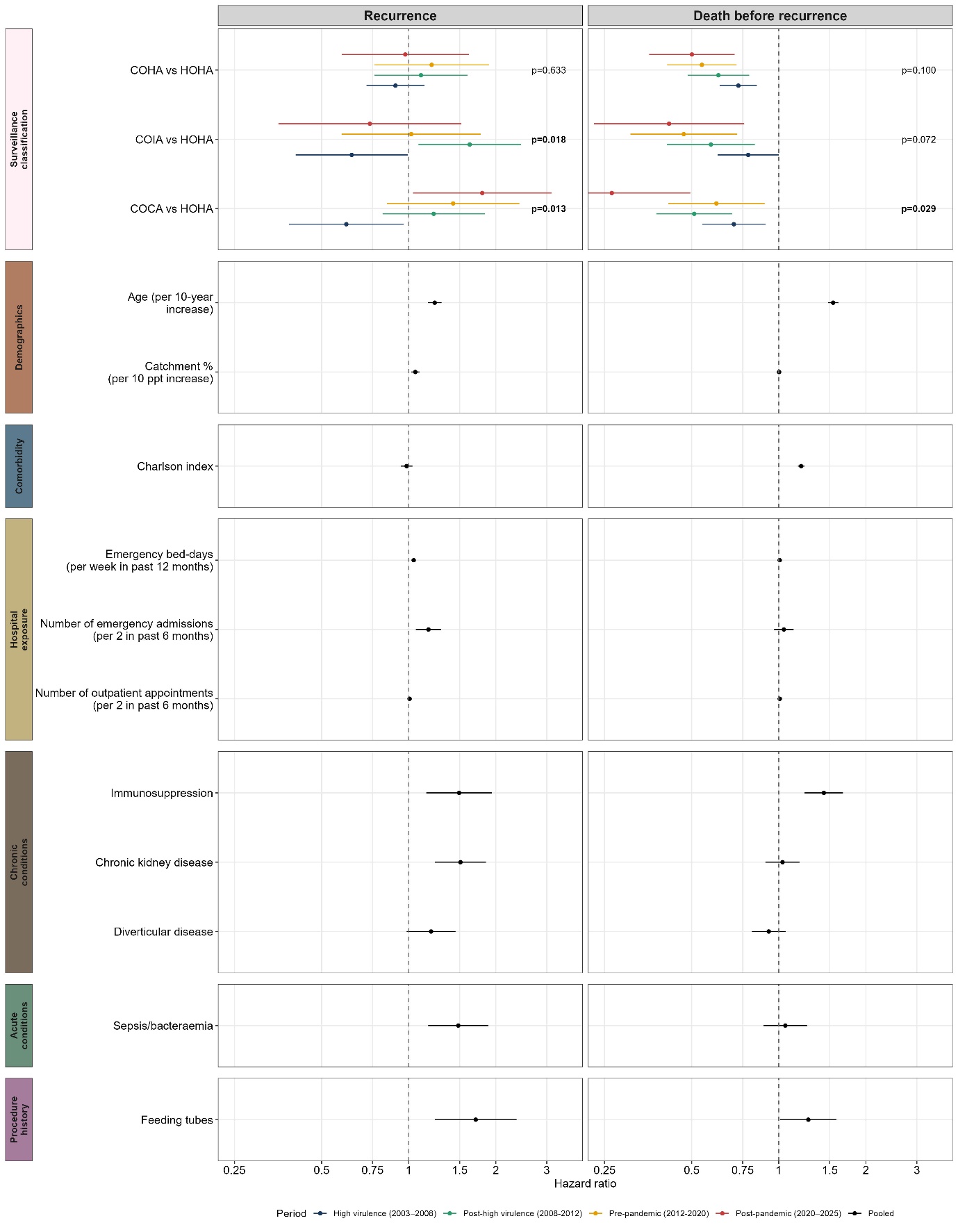

Note: Overall heterogeneity p-values for surveillance classification were 0.010 (recurrence) and 0.029 (death). Models were stratified by period, and robust standard errors were clustered by patient. Plot truncated at [0.25, 3]. The lower 95% CI for “post-pandemic” COCA vs HOHA (death) is 0.14.

**Supplementary Figure S16. Risk factors for recurrence and death before recurrence, also adjusting for blood test results**

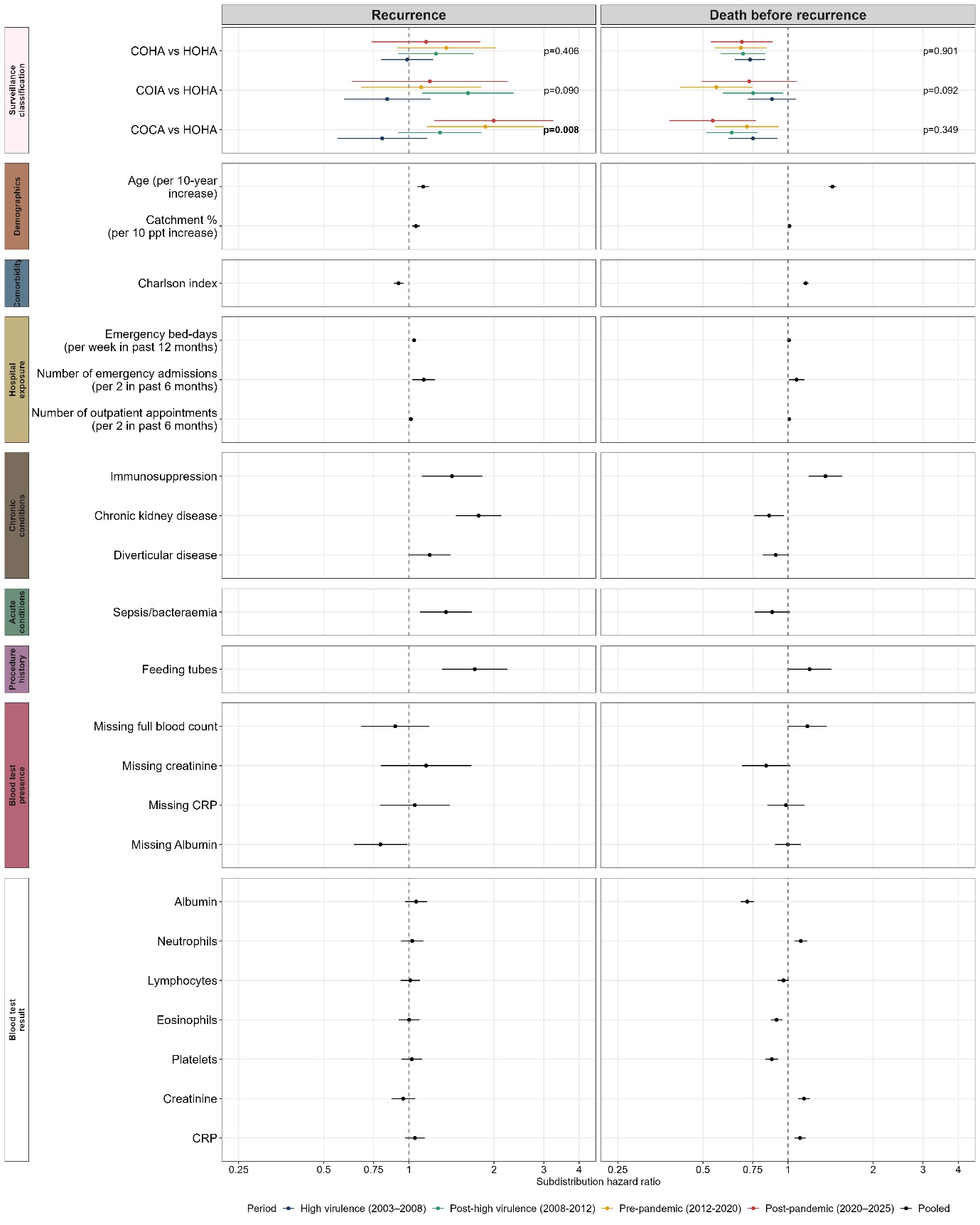

Note: Overall heterogeneity p-values for surveillance classification were 0.022 (recurrence) and 0.333 (death). Models were stratified by period, and robust standard errors were clustered by patient. Haemoglobin was excluded from models due to evidence of an interaction effect with albumin on mortality influencing estimates. Replacing albumin with haemoglobin did not change effects (no significant association with recurrence or death before recurrence seen for haemoglobin).

**Supplementary Figure S17. Cause-specific risk factors for recurrence and death before recurrence, also adjusting for blood test results**

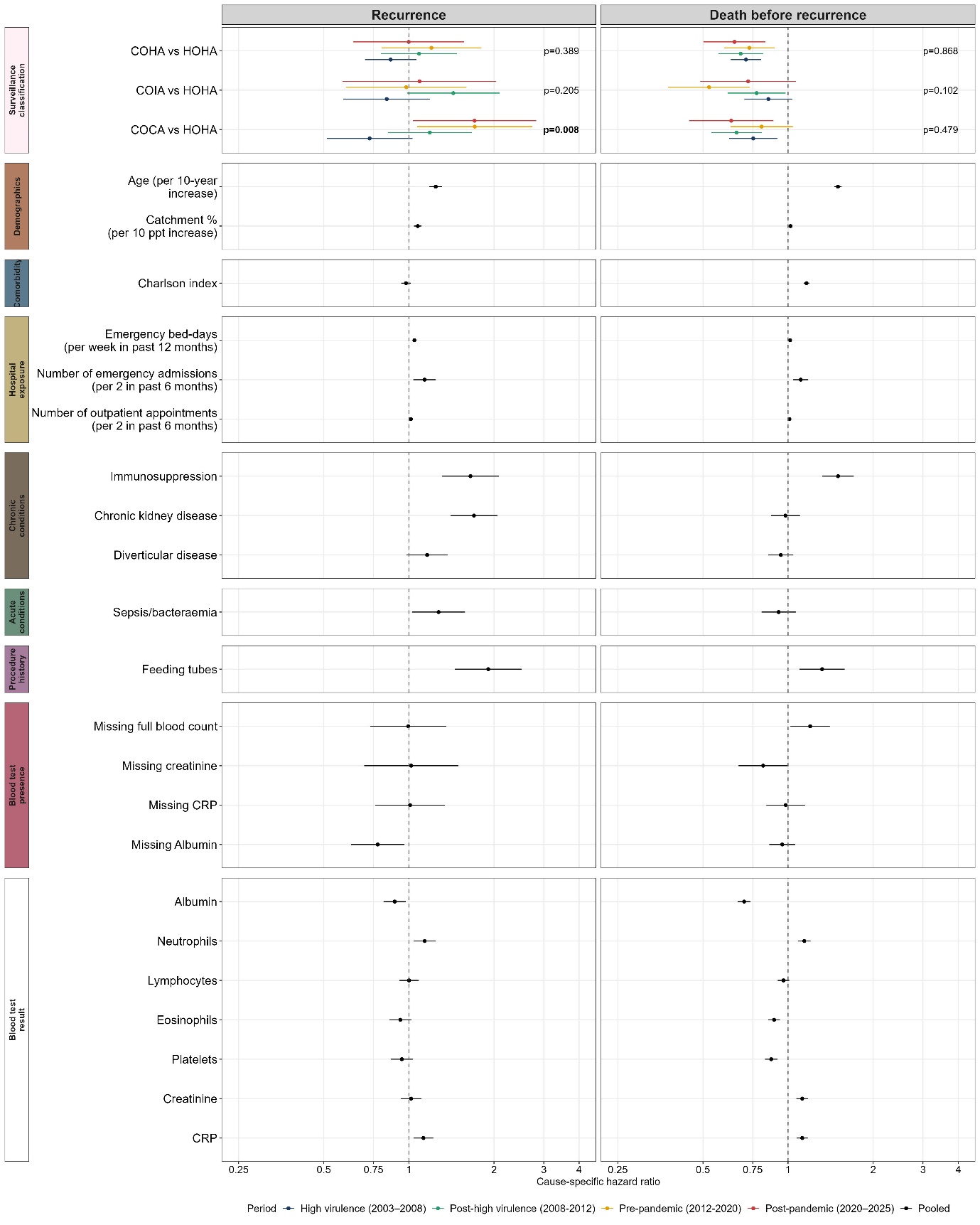

**Supplementary Figure S18. Risk factors for recurrence and death before recurrence, also adjusting for additional admission-related variables**

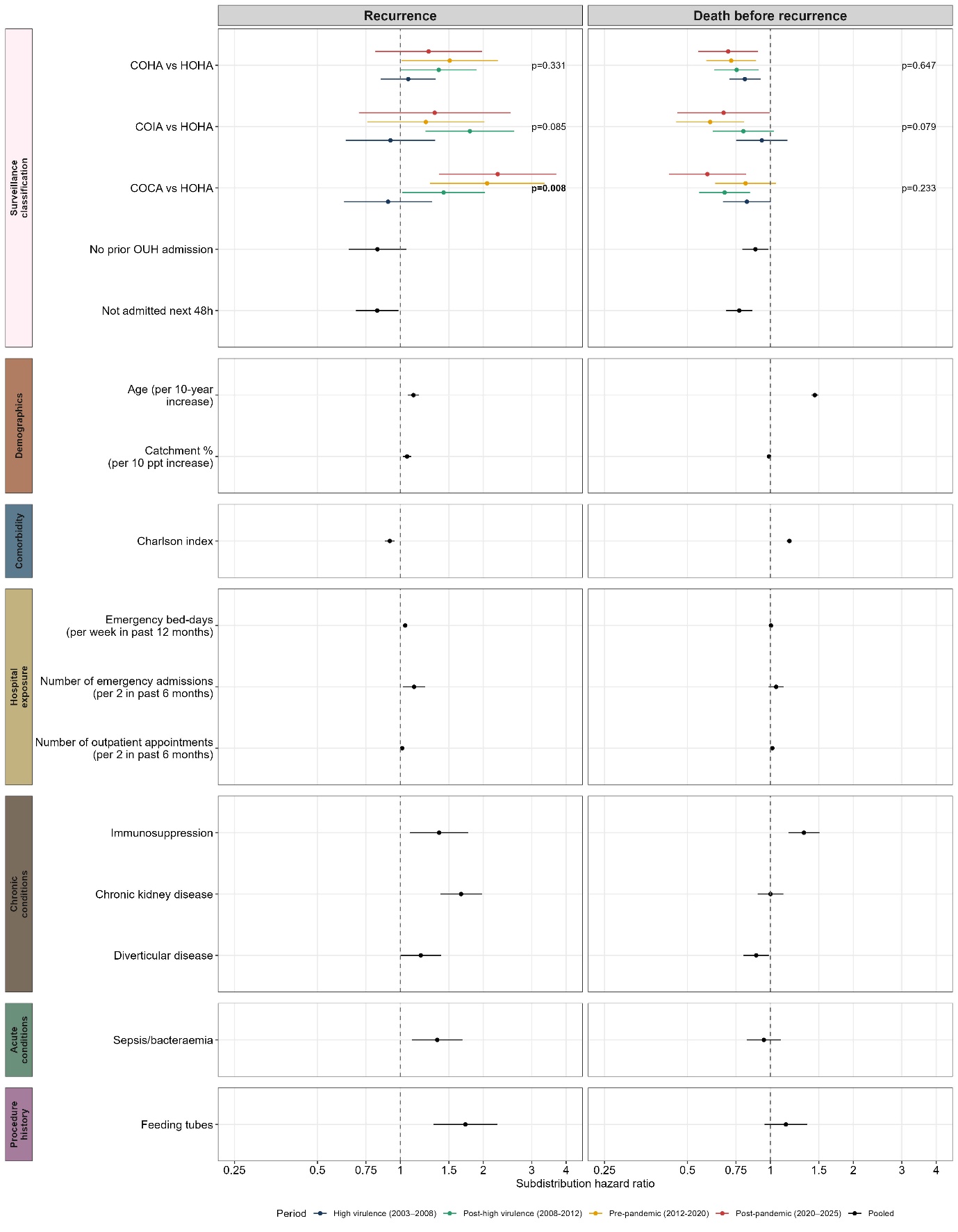

**Supplementary Figure S19. Percentage of samples by location at the time of sample collection over calendar time by surveillance classification**
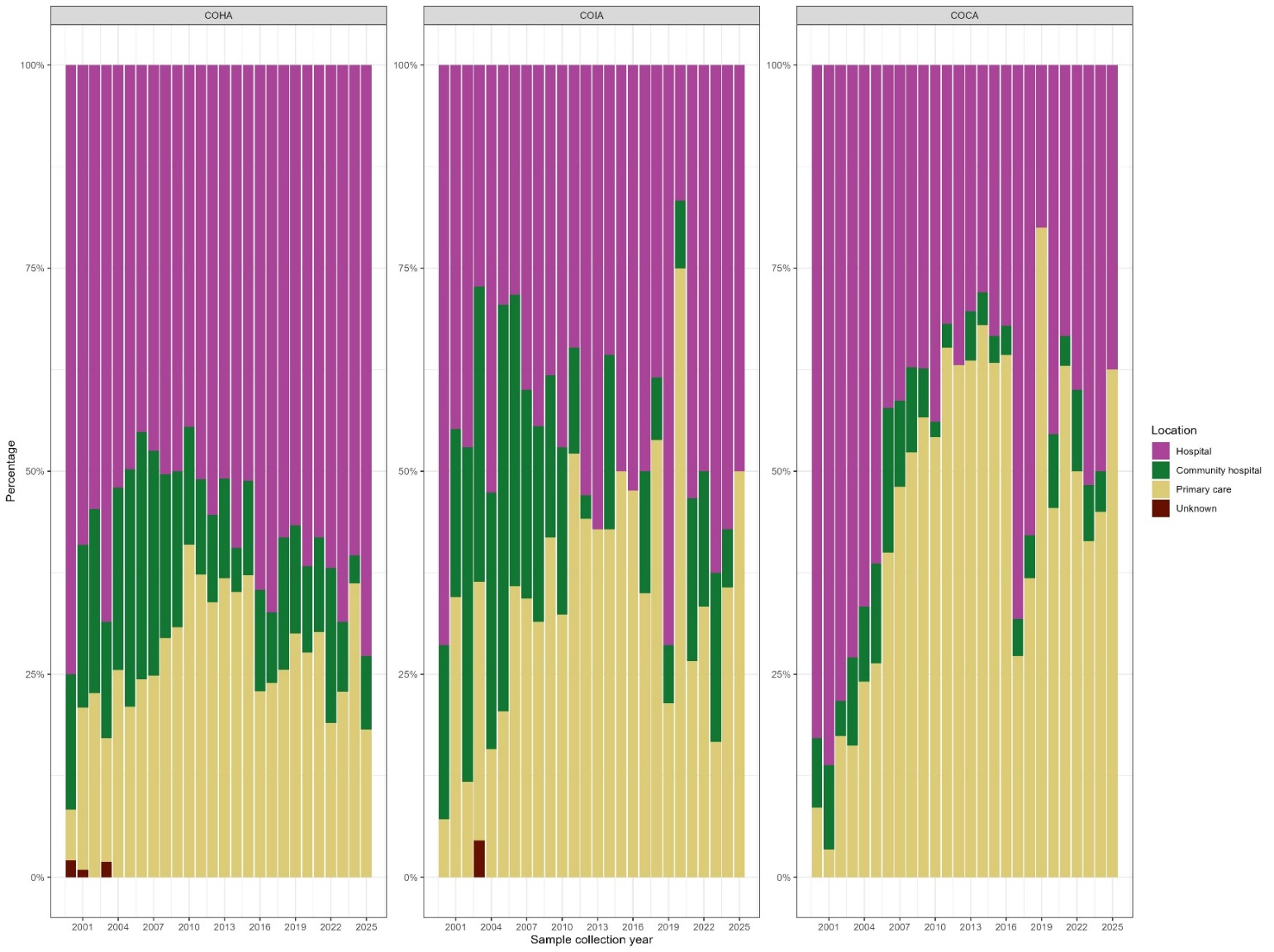
