## Supplementary methods for "Increasing risks of recurrence following community-associated *Clostridioides difficile* infection in Oxfordshire 2000-2025"

Definitions of factors studied**

| **Category** | **Factor** | **Definition** | **Truncation** |
| --- | --- | --- | --- |
| Surveillance classification | Surveillance classification (HOHA, COHA, COIA, COCA) | Hospital-onset hospital-associated (HOHA) samples were collected >48h from admission^1^. Community-onset hospital-associated (COHA) and community-onset indeterminate-association (COIA) samples were collected in the community or within ≤48h of admission, from patients last discharged within ≤4 or 4-≤12 weeks, respectively. Community-onset community-associated (COCA) samples were collected in the community or within ≤48h of admission, from patients not hospitalised in the past 12 weeks. |  |
| Demographics | Age | Patient age at time of sample collection | 25-95 years (approximate 1^st^-99^th^ percentiles) |
|  | Sex | Patient sex |  |
|  | Deprivation | Index of Multiple Derivation (IMD)^2^ of the LSOA of most recent patient address within the past 10 years (or the following five years if no match was found). Deprivation was measured by IMD percentile where lower ranks/percentiles indicate higher deprivation, but effects in models were displayed to show effect of increased deprivation. Addresses were matched to the different IMD indices as follows: observations prior to April 2007 to the 2004 indices, observations April 2007 – March 2010 to 2007 indices, observations April 2010 to March 2015 to 2010 indices, observations April 2015 – March 2019 to 2015 indices, observations April 2019 onwards to 2019 indices. |  |
|  | Catchment | Catchment percentage of the MSOA of the most recent patient address within the past 10 years (or the following five years if no match was found). Catchment percentage (proportion of residents in a MSOA admitted to OUH versus other hospitals (trusts) was obtained from the Office for Health Improvement and Disparities^3,4^ and matched to addresses by year. Catchment data was available for 2011-2020 and 2023-2024; address observations pre-2011 were assigned the 2011 value, observations 2021/2022 the 2023 value, and observations after 2024 the 2024 value. |  |
| Prior testing | Prior negative test result | Prior ELISA-Negative test result (before April 2012) or GDH-Negative test result (April 2012 onwards) in the past 6 months before sample collection for current CDI |  |
|  | Prior carriage | Prior GDH-Positive, Toxin-ELISA-Negative test result (April 2012 onwards) in the past 6 months before sample collection for current CDI |  |
|  | Prior CDI | Prior ELISA-Positive test result (before April 2012) or GDH-Positive, Toxin-ELISA-Positive test result (April 2012 onwards) in the past 6 months before sample collection for current CDI |  |
| Comorbidity scores | Charlson | Charlson comorbidity index using Quan weights^5^, calculated with the R comorbidity package^6^, using secondary codes from the diagnostic-dominant episode of the current spell for hospitalised patients^7^, plus a one-year lookback including all ICD-10 codes recorded during inpatient admissions, outpatient attendances, and A&E attendances (2020 onwards). |  |
|  | Elixhauser | Elixhauser comorbidity index using Swiss weights^8^, calculated with the R comorbidity package^6^, using secondary codes from the diagnostic-dominant episode of the current spell for hospitalised patients^7^, plus a one-year lookback including all ICD-10 codes recorded during inpatient admissions, outpatient attendances, and A&E attendances (2020 onwards). |  |
|  | Gilbert | Hospital Frailty Risk Score calculated with all codes from hospital episodes ending before sample collection, plus prior codes from outpatient attendances and A&E attendances (2020 onwards), using a two-year lookback^9^ |  |
| Hospital exposures | Emergency bed-days | Number of days in the past 12 months where the patient was admitted under an emergency hospital spell at OUH, excluding any current admission. | Individual spells truncated at 120 days to avoid influence from data entry errors and outliers |
|  | Elective bed-days | Number of days in the past 12 months where the patient was admitted under an elective hospital spell at OUH, excluding any current admission. | Individual spells truncated at 120 days to avoid influence from data entry errors and outliers |
|  | Number of emergency admissions | Number of emergency hospital admissions at OUH in the past 6 months |  |
|  | Number of outpatient appointments | Number of attended outpatient appointments at OUH in the past 6 months, counting multiple appointments on the same day separately |  |
|  | Recent surgical admission | Any admission under a surgical treatment function in the past 3 months (100-111, 115, 120, 130, 140, 144, 145, 150, 160, 170-173, 190). |  |
|  | Recent GI admission | Any admission under a GI treatment function in the past 3 months (104-106, 251, 301, 306) |  |
| Chronic conditions (Any ICD-10 code in the past 5 years, excluding codes from the current admission for hospitalised patients) | Palliative care | Code Z515 following Yoon et al.^10^ |  |
|  | Immunosuppression | Following Yoon et al.^10^, codes indicating metastatic cancer, haematological malignancies, primary immunodeficiencies, and end-stage liver disease, but excluding AIDS/HIV on clinical advice. |  |
|  | Dementia | Including Alzheimer’s disease, vascular and unspecified dementia, and senility as categorised by Gilbert et al.^9^. |  |
|  | Anaemia | Codes D50-D61 and D63-D64, excluding acute post-haemorrhagic anaemia |  |
|  | Cancer | Codes from the Summary Hospital-level Mortality Indicator (SHMI)^11^ diagnosis cluster “Cancer”, excluding unspecified and benign neoplasms and non-malignant breast conditions |  |
|  | Chronic kidney disease | Codes from the SHMI diagnosis group 100 |  |
|  | Heart failure | Codes from the SHMI diagnosis group 65 |  |
|  | Hypertension | Codes from the SHMI diagnosis group 56 |  |
|  | Other heart disease | Remaining codes from the SHMI diagnosis cluster “Heart”, excluding heart failure, hypertension, and acute MI. Diagnosis groups 54-55, 58-64. |  |
|  | COPD | Codes from the SHMI diagnosis group 75 |  |
|  | IBD | Codes from the SHMI diagnosis group 88 |  |
|  | Diverticular disease | Codes from the SHMI diagnosis group 90 |  |
|  | Oesophegal disease | Codes from the SHMI diagnosis group 84 |  |
|  | Cerebrovascular disease | Codes from the SHMI diagnosis group 67 |  |
|  | Liver disease | Codes from the SHMI diagnosis cluster “Liver”, diagnosis groups 92-94 |  |
|  | Diabetes | Codes from the SHMI diagnosis groups 34 and 35 (diabetes with and without complications) |  |
| Acute conditions (Any ICD-10 code in the past 90 days, excluding codes from the current admission for hospitalised patients) | Acute renal failure | Codes from the SHMI diagnosis group 99 |  |
|  | Pneumonia/bronchitis | Codes from the SHMI diagnosis groups 73 and 74 |  |
|  | Sepsis/bacteraemia | Codes from the SHMI diagnosis group 2 (septicaemia (except in labour), shock). |  |
|  | UTI | Codes from the SHMI diagnosis group 101 |  |
| Procedure history (Any OPCS-4 code in the past 5 years, excluding codes from the current admission for hospitalised patients) | Transplant | Transplant history following the National Health Service Digital code list^12^ |  |
|  | Feeding tubes | Codes for enteral feeding tube insertion, management, and closure (G083, G383, G341-G343, G345, G445, G447, G475, G675, G601-G604) |  |
| Blood test result (closest result within [-24, +48h] of sample collection) | Neutrophils | Pooled median (standard deviation) and truncation bounds:  9.7 (6.8), 0-27.3 | Missing values were set to the pooled median across periods and a binary indicator included for missing full blood count (FBC), creatinine or C-reactive protein (CRP). If any full blood count (FBC) component was missing, all FBC components were set to their pooled medians, as they are routinely measured together. Values were truncated at 95^th^ (pooled) percentiles if naturally containing zero, otherwise 2.5^th^ – 97.5^th^ percentiles to reduce outlier influence. For comparability, blood tests were standardised so that a one-unit increase represented a global standard deviation. |
|  | Eosinophils | Pooled median (standard deviation) and truncation bounds: 0.09 (0.13), 0-0.45 |  |
|  | Lymphocytes | Pooled median (standard deviation) and truncation bounds: 1.1 (0.7), 0-2.8 |  |
|  | Haemoglobin | Pooled median (standard deviation) and truncation bounds: 109 (18.7), 75-151 |  |
|  | Platelets | Pooled median (standard deviation) and truncation bounds: 298 (140), 2-598 |  |
|  | CRP | Pooled median (standard deviation) and truncation bounds: 96 (70), 0.1-254 |  |
|  | Creatinine | Pooled median (standard deviation) and truncation bounds: 89 (107), 42-538 |  |
|  | Albumin | Pooled median (standard deviation) and truncation bounds:  25.1 (6.2), 11.9-37.4 |  |

**Blood test adjustments**
**Albumin**
The method of measuring albumin changed in January 2015 to a more sensitive Bromocresol Purple (BCP) method from the previous Bromocresol Green (BCG) assay principle. Albumin observations before January 2015 were therefore adjusted using the formula 1.0192*albumin – 8.4921, following results from a validation report on the new method. Before this adjustment, albumin observations were truncated to 15-55, based on the “clinically relevant” range of values the validation procedure was performed on. This was equivalent to the approximate 1-99^th^ percentiles of the raw data.

**Creatinine**
There were two changes to methods used to analyse blood creatinine. In August 2009, there was a change from an international initiative to align creatinine values from all methods to a reference isotype dilution mass spectrometry. In January 2015, there was a change of method from a traditional Jaffe method to a more specific enzymatic method. In order to estimate adjustments from these changes, we used all creatinine results three months before and after each change from patients tested for CDI (positive and negative results), with each creatinine observation matched to a CDI test and all CDI tests de-duplicated by 28 days**.** Linear regression was used to estimate a linear transformation between pre-change and post-change deciles, yielding the following formulae:

*Creatinine_After2009Change = 0.9508 * creatinine – 10.5709
Creatinine_After2015Change = 1.0249 * creatinine + 8.0059*

These corrections were applied sequentially to the data. Before the first correction, creatinine values were truncated to 31-706 (approximate 1-99^th^ percentiles). Both albumin and creatinine were further truncated in models, as with other blood result variables.

**Sensitivity analysis: calendar time as a continuous variable**
To try to estimate when changes in recurrence risk occurred more precisely, we also considered an over-parameterised generalised additive model (GAM) for recurrence over the full study period, modelling continuous calendar time by surveillance classification with penalised thin plate splines (*mgcv* package^13^ with 50 basis functions). This model was fit on modified survival data that allowed for an approximation of the Fine-Gray subdistribution hazards using a Cox specification. Specifically, individuals who experienced the competing event (death) were retained in the risk set until the administrative censoring date (March 16^th^ 2025), following the “censoring-complete” formulation of Fine and Gray^14^.
